# Weeding Through the Haze: A Survey on Cannabis Use Among People Living with Parkinson’s Disease in the US

**DOI:** 10.1101/2020.06.24.20139162

**Authors:** Megan P. Feeney, Danny Bega, Benzi M. Kluger, A. Jon Stoessl, Christiana M. Evers, Rebeca De Leon, James C. Beck

## Abstract

Symptomatic management of Parkinson’s disease (PD) is complex and many symptoms, especially non-motor symptoms, are not effectively addressed with current medications. In the US, cannabis has become more widely available for medical and recreational use, permitting those in the PD community to try alternative means of symptom control. However, little is known about the attitudes towards, and experiences with, cannabis use among those living with PD. To address this shortcoming, we distributed an anonymous survey to 7,607 people with PD in January 2020 and received 1,339 responses (17.6%). 1,064 complete responses were available for analysis. Respondents represented 49 states with a mean age of 71.2 years (± 8.3) and mean PD duration of 7.4 years (± 6.2). About a quarter of respondents (24.5%) reported cannabis use within the previous six months. Age and gender were found to be predictors of cannabis use in this sample (Age OR = 0.95, 95% CI 0.93 to 0.97; Male OR = 1.44, 95% CI 1.03 to 2.03). Users reported learning about cannabis use from the internet/news (30.5%) and friends or other people with PD (26.0%). Cannabis users were more likely to report insufficient control of their non-motor symptoms with prescription medications than non-users (p = 0.03). Cannabis was primarily used for PD (63.6%) and was most often used to treat nonmotor symptoms of anxiety (45.5%), pain (44.0%), and sleep disorders (44.0%). However, nearly a quarter of users (23.0%) also reported they had stopped cannabis use in the previous six months, primarily due to a lack of symptom improvement (35.5%). Three quarters of respondents (75.5%) did not use cannabis, primarily because there was a lack of scientific evidence supporting efficacy (59.9%). Our results suggest that the lack of formal guidance or research evidence about cannabis for PD may in part underlie inconsistencies in both use and reported effectiveness.

## Introduction

Parkinson’s disease (PD) is the second most common neurodegenerative disorder, affecting more than 1 million Americans at a cost to society of more than $50 Billion dollars.^1,2^ While PD is typically defined clinically by four cardinal motor symptoms that include resting tremor, bradykinesia, rigidity and postural instability,^3^ the symptoms of PD are broader. Non-motor manifestations of PD are varied and include sleep disturbances such as insomnia, sleep fragmentation and REM sleep behavior disorder (RBD), cognitive changes, depression, anxiety, hallucinations, hyposmia, pain and autonomic dysfunction (constipation, orthostatic hypotension and urinary incontinence).^4,5^ Additionally, treatment of PD with levodopa or other dopaminergic agents is commonly associated with involuntary movements (levodopa-induced dyskinesias, LID) that may be troublesome for some patients. The efficacious control of LID and of non-motor features represents a major therapeutic challenge that is often unmet for those living with PD.^5^ Therefore, many of those living with PD in the US are interested in complementary and alternative or integrative therapies (CAIM) that may bring relief to their troubling non-motor symptoms.^6,7^

The medicinal use of cannabis represents a novel, alternative approach toward PD symptom control. Preclinical evidence suggests that cannabinoids could be widely beneficial to neurodegenerative diseases, including PD.^8,9^ A small survey in Colorado of individuals with PD found that the handful of identified cannabis users reported it to be among the most effective of complementary therapies.^10,11^ However, there remains a lack of evidence of both efficacy and safety to support the recommendation of the use of cannabis as a therapy for PD.^12,13^

Nevertheless, there is an increasing interest in the use of medical cannabis. In one recent survey, 95% of movement disorder specialist neurologists reported being asked to prescribe medical cannabis to their patients.^14^ As of October, 2020, 47 of 50 states in the US permitted the sale use, or consumption of hemp-sourced cannabidiol (CBD) products containing less than 0.3% tetrahydrocannabinol (THC), 33 of 50 states permitted the use of marijuana-sourced products for qualifying medicinal purposes and 11 of 50 states permitted recreational use of marijuana-sourced products (Fig. 1).^12,15–17^ As a result, broad numbers of individuals living with PD now have access to cannabis, with and without a physician’s recommendation. Adding to this mix is the publicity that surrounds cannabis use for PD.^18^ In this study, we administered an anonymous survey to Parkinson’s Foundation constituents with PD, with the goal of understanding the motivations and experiences of cannabis use among people living with PD. Given the wide range of cannabis available to people with PD within the US, this study aimed to capture community viewpoints, use and self-perceived outcomes. This study did not attempt to assess or determine the efficacy of specific types or formulations of cannabis (CBD vs. THC, for example).

**Figure 1:**
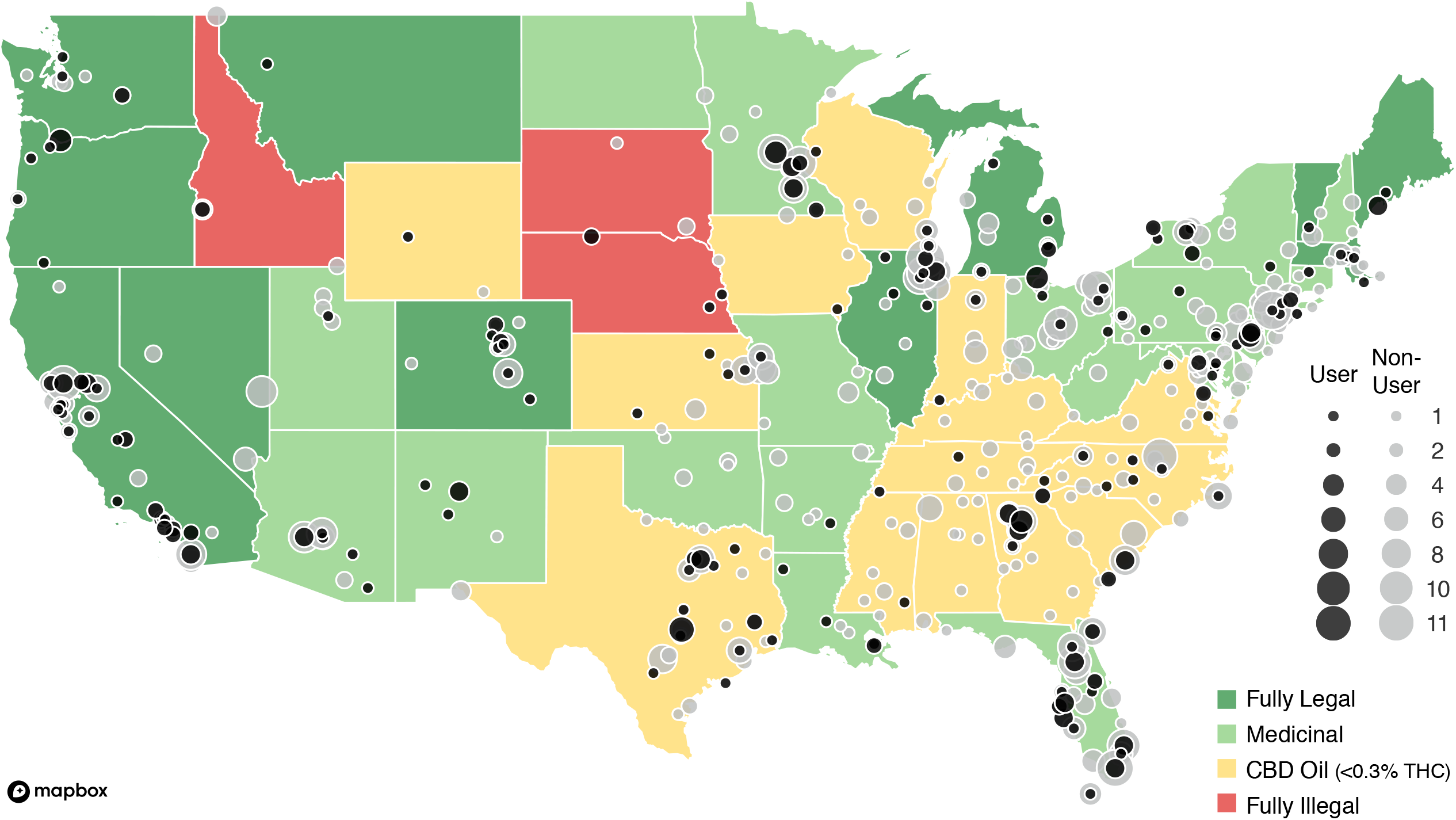
A Geographic representation of survey participants. Color of state depicts the legality of cannabis as of October 2020. Black dots represent respondents who identified as cannabis users and grey dots represent respondents who identified as non-users. Dot size corresponds to number of respondents by zip code. Rhode Island was the only state without survey respondents. Outline of US from Mapbox.

## Results

### Survey population

An anonymous, electronic survey (see Supplementary Information) was sent by email on January 17, 2020 to 7,607 individuals with PD who had previously engaged with the Parkinson’s Foundation, primarily through attendance of in-person or online educational event or calls to the Foundation’s toll-free Helpline. When the survey closed on February 7, 2020, 1339 surveys had been returned, providing a response rate of 17.6%. Of the returned surveys,1064 individuals from 49 states had provided complete responses. State representation is shown in Fig. 1. Demographic information about survey respondents can be found in Table 1. Briefly, respondents averaged 71.2 years (± 8.3) in age with a mean PD duration of 7.4 years (± 6.2). Most (68.5%) respondents held a bachelor’s degree or higher and were currently retired or not employed (87.3%). However, women completing the survey were on average 1.3 years younger than men (70.5 ± 8.5 vs 71.8 ± 8.2 years; p = 0.02).

**Table 1:** Characteristics of the Study Population

|  |  |
| --- | --- |
| Age, mean (SD) | 71.2 (8.3) |
| Duration of disease, mean (SD) | 7.44 (6.2) |
| Male, n (%) | 559 (52.5) |
| Female, n (%) | 500 (47.0) |
| Other, n (%) | 1 (0.1) |
| Prefer not to say, n (%) | 4 (0.4) |
| Race |  |
| White, n (%) | 977 (91.8) |
| Hispanic, n (%) | 25 (2.3) |
| African American, n (%) | 16 (1.5) |
| Asian, n (%) | 11 (1.0) |
| American Indian or Alaska Native, n (%) | 3 (0.3) |
| Mixed, n (%) | 7 (0.7) |
| Some other race, ethnicity or origin, n (%) | 5 (0.5) |
| Prefer not to answer, n (%) | 20 (1.9) |
| Education level |  |
| Less than high school, n (%) | 2 (0.2) |
| High school, n (%) | 81 (7.6) |
| Some College, n (%) | 166 (15.6) |
| Trade/Technical, n (%) | 28 (2.6) |
| Associate's, n (%) | 58 (5.5) |
| Bachelor's, n (%) | 331 (31.1) |
| Master's, n (%) | 260 (24.4) |
| Professional, n (%) | 78 (7.3) |
| Doctorate, n (%) | 60 (5.6) |
| Income |  |
| < \$25,000, n (%) | 75 (7.0) |
| \$25,000 - \$34,999, n (%) | 70 (6.6) |
| \$35,000 - \$49,999, n (%) | 109 (10.2) |
| \$50,000 - \$74,999, n (%) | 193 (18.1) |
| \$75,000 - \$99,999, n (%) | 141 (13.3) |
| \$100,000 - \$149,999, n (%) | 145 (13.6) |
| \$150,000 - \$199,999, n (%) | 53 (5.0) |
| > \$200,000, n (%) | 59 (5.5) |
| Prefer not to answer, n (%) | 219 (20.6) |
| Marital Status |  |
| Single, n (%) | 44 (4.1) |
| Married or DP, n (%) | 809 (76.0) |
| Separated, n (%) | 6 (0.6) |
| Divorced, n (%) | 123 (11.6) |
| Widowed, n (%) | 82 (7.7) |
| Employment |  |
| Full-Time, n (%) | 69 (6.5) |
| Part-Time, n (%) | 30 (2.8) |
| Self-Employed, n (%) | 36 (3.4) |
| Unemployed, n (%) | 18 (1.7) |
| Retired, n (%) | 834 (78.4) |
| Unable to work, n (%) | 77 (7.2) |
| Care Partner |  |
| No, n (%) | 267 (25.1) |
| Yes, n (%) | 797 (74.9) |
| Total Participation: 1,064 |  |

SD Standard deviation

### Non-Users are the majority

We began our survey by asking participants if they had used marijuana or cannabis in the last six months. Most respondents (75.5%, 803/1064) did not report cannabis use within this time frame. Those individuals who identified as non-users were then asked to select up to three reasons for not using cannabis (see Fig. 2). The most frequently identified reasons for non-use included a lack of evidence of the efficacy of cannabis (59.9%, 481/803), a fear of cannabis side effects (34.9%, 280/803) and “other reasons” (31.3%, 250/803), which led to an open text field for respondents to provide additional information. The theme of these open text responses focused on the legality of cannabis (both state and federal), a lack of a need to use cannabis, as well as a disinterest in cannabis overall.

**Figure 2:**
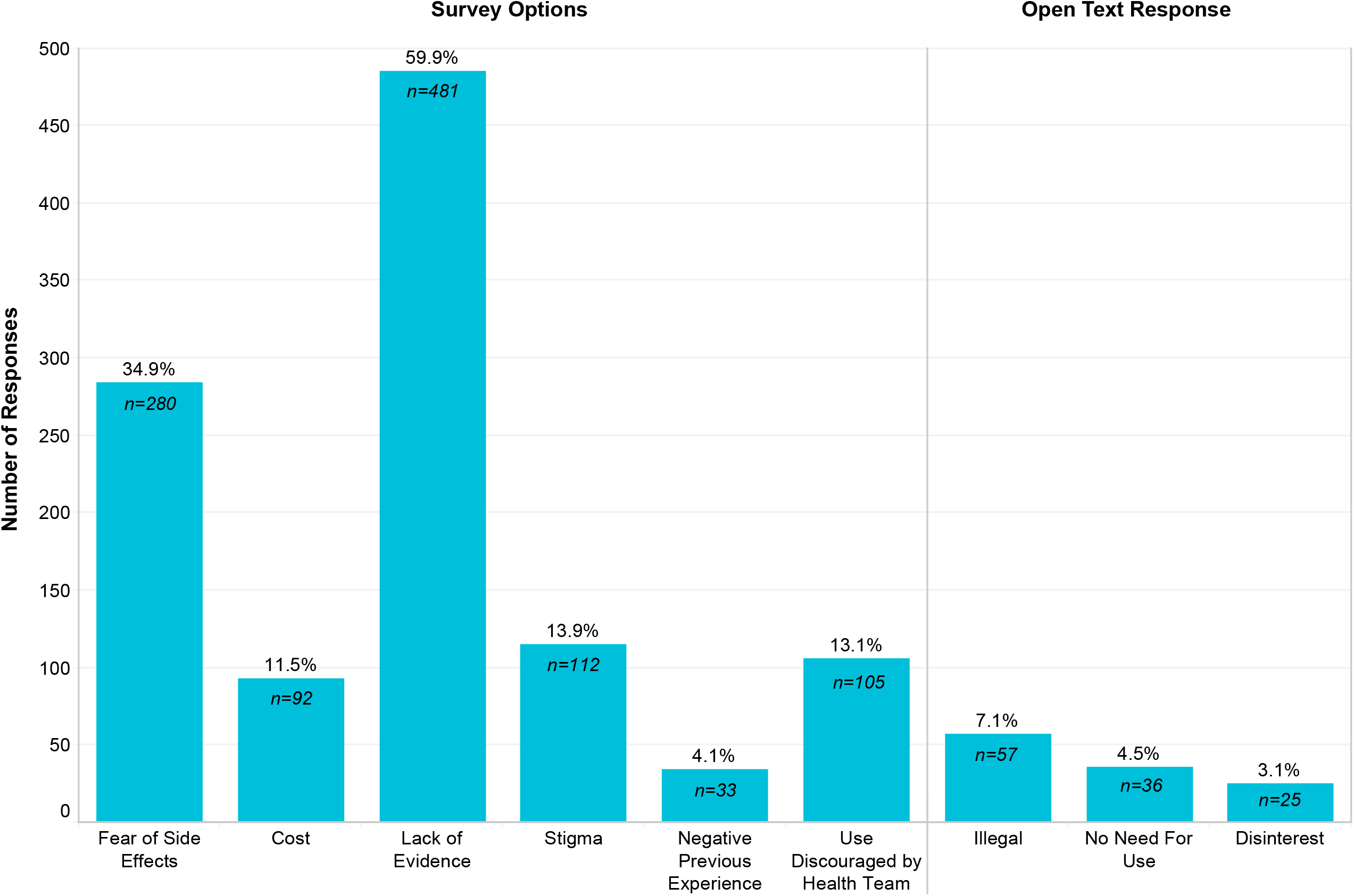
Primary reasons for not using cannabis in the past 6 months (n=803). Responses were either pre-identified in the survey as options listed in the category labels or individuals selected “other” and their open text response was analyzed thematically.

We then asked non-users to select up to three reasons that might influence them to consider using marijuana or cannabis. The majority of non-users (76.0%, 610/803) indicated that evidence which supported cannabis use for PD would be most influential in any deliberations to begin cannabis use. Other factors that would also be important in their decision would be if their health team encouraged its use (54.6%, 438/803) or if cannabis side effects were better understood (35.5%, 285/803).

### Motivations and Routines Around Cannabis Use

Overall, a quarter of respondents (24.5%, 261/1064) reported they had used cannabis within the previous six months. Age and gender were found to be predictors of cannabis use in this sample (Age OR = 0.95, 95% CI 0.93 to 0.97; Male OR = 1.44, 95% CI 1.03 to 2.03; see Table 2). Cannabis users were younger by an average of 3 years in age than non-users (69 ± 8.6 vs 72 ± 8.1 years; p < 0.001). When respondents were asked why they used cannabis, the two most common reasons selected were for medical reasons related to their PD only (63.6%, 166/261) or for the combination of both medical and nonmedical (e.g., recreational) reasons (21.5%, 56/261; see Table 3). Age and disease duration were predictors of cannabis use for PD reasons; however, significance disappeared when a provider referral was included in the regression model. For those who used cannabis explicitly for the PD or for both medical and non-medical reasons, about half (46.8%,104/222) indicated they used cannabis for PD symptoms in general and a similar number (43.2%, 96/222) indicated they used cannabis for specific PD symptoms (see Table 3). Comparable findings were identified through thematic analysis of open-text responses. Among the 246 users (85.1%) who reported their cannabis use was for any medical reason or for both medical and non-medical reasons, the most commonly identified reasons for trying cannabis included curiosity (22.8%, 56/246), the idea that cannabis was a natural substance (16.7%, 41/246) and word of mouth (11.4%, 28/246).

**Table 2:** Adjusted Analysis of Cannabis Use

Multivariable Logistic Regression
| Predictors of Cannabis Use | OR (95% CI) | P |
| --- | --- | --- |
| Marital Status: Married/DP | 1.02 (0.59-1.76) | 0.95 |
| Gender: Male | 1.44 (1.03-2.03) | 0.04* |
| Race: White | 1.03 (0.56-2.00) | 0.93 |
| Education: ≥ Bachelors | .75 (0.53-1.07) | 0.12 |
| Income: ≥ \$100,000 | .80 (0.54-1.18) | 0.27 |
| Employment: Not working | 1.36 (0.82-2.31) | 0.25 |
| Age | 0.95 (0.93-0.97) | <0.00* |
| Disease Duration | 1.00 (0.98-1.03) | 0.74 |
| Primary Health Concern: Yes | 1.00 (0.59-1.75) | 1.00 |
| Care Partner: Yes | .95 (0.57-1.60) | 0.85 |
| Medications: None | 1.56 (0.71-3.26) | 0.24 |

**OR**, Odds Ratio; **CI**, Confidence Interval.

**Table 3:**
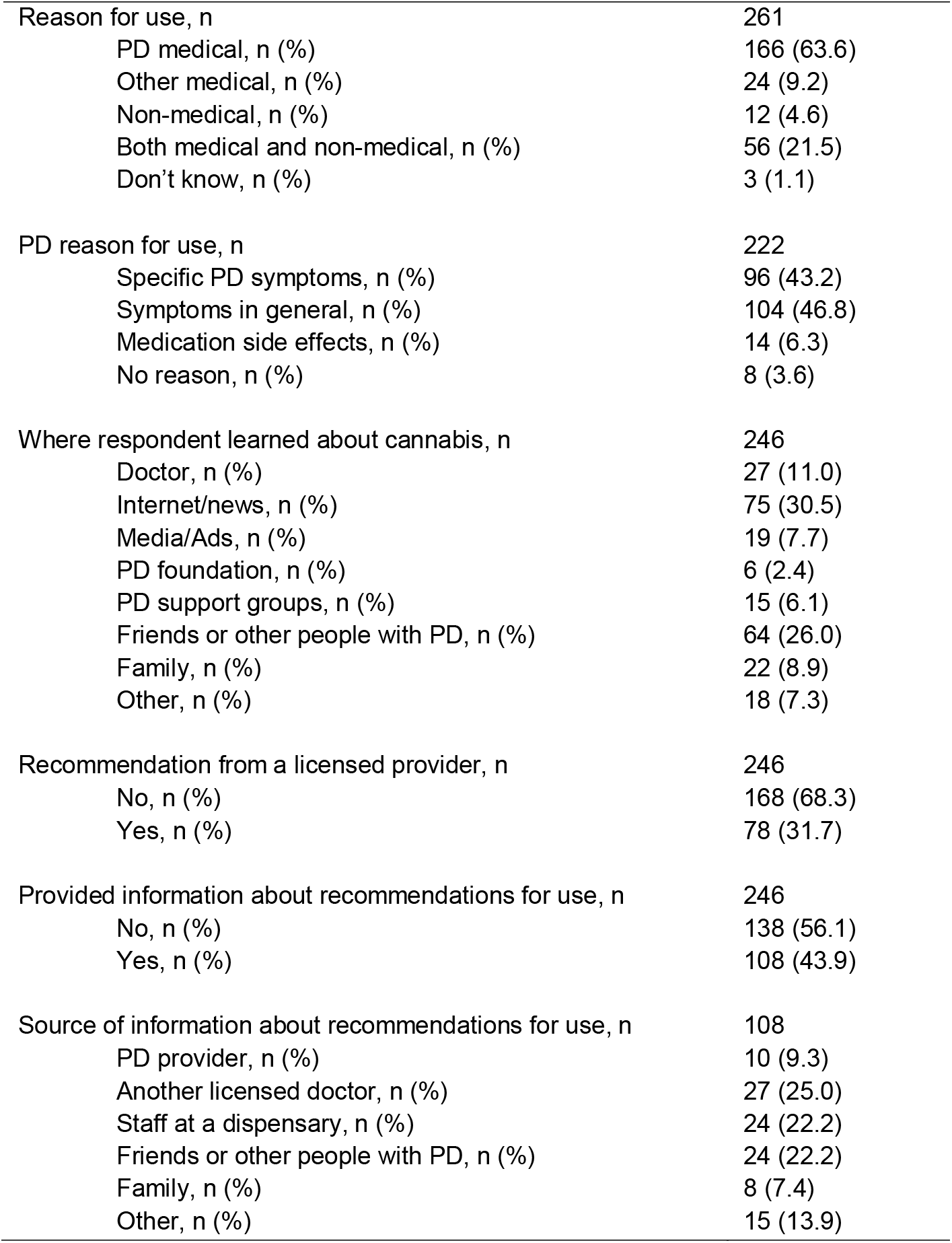
Reasons for Cannabis Use

We next sought to understand how cannabis users acquired their knowledge about cannabis as a potential therapy (see Table 3). For the majority of users who used cannabis for any medical reason or for both medical and non-medical reasons (excluding recreational users and users who did not know their reason for use), the most common sources of information were from the internet/news (30.5%, 75/246) and from friends or other people with PD (26.0%, 64/246). Most of these users (64.4%, 168/246) had not received a recommendation to use cannabis from a licensed doctor or provider. However, receiving a recommendation from a licensed provider was associated with younger age (OR = 0.95, 95% CI 0.91 to 0.99).

Following up our question on knowledge, we asked these medical users and medical and non-medical users how they learned how to use cannabis (such as dosage, type and frequency of use). We found that a majority of users (56.1%, 138/246) reported that they were not provided any information about recommendations on how to use cannabis. Of the users who were provided information (43.9%, 108/246), the most common sources of information were reported to be a non-PD doctor (25.0%, 27/108), staff at a dispensary (22.2%, 24/108) and friends or other people with PD (22.2%, 24/108).

Among all users, we also sought to better understand the cannabis formulations, strengths and routes of administration they most often used. Two-thirds (64.4%, 168/261) considered themselves “as needed” users and the remainder (35.6%, 93/261) considered themselves “regular” users. Being male, identifying PD as the primary health concern, a shorter disease duration and having been provided a recommendation were all associated with “regular” cannabis use (see Table 4). The daily routine of cannabis use varied and a summary can be found in Table 5. The majority of users reported their time of use at the end of the day, either in the evening (25.3%, 66/261) or at bedtime (26.4%, 69/261, see Fig. 3A).

**Table 4:** Adjusted Analysis of “Regular” Cannabis Use

Multivariable Logistic Regression
| Predictors of “Regular” Cannabis Use | OR (95% CI) | P |
| --- | --- | --- |
| Marital Status: Married/DP | 0.91 (0.28-3.10) | 0.88 |
| Gender: Male | 2.39 (1.21-4.85) | 0.01* |
| Race: White | 2.43 (0.70-10.09) | 0.18 |
| Education: ≥ Bachelors | .80 (0.39-1.62) | 0.53 |
| Income: ≥ \$100,000 | 1.14 (0.51-2.52) | 0.75 |
| Employment: Not working | 1.79 (0.62-5.44) | 0.29 |
| Age | 0.99 (0.95-1.03) | 0.58 |
| Disease Duration | 0.94 (0.88-0.99) | 0.03* |
| Primary Health Concern: Yes | 5.97 (1.49-41.18) | 0.03* |
| Care Partner: Yes | 0.48 (0.15-1.47) | 0.20 |
| Received Recommendation: Yes | 2.90 (1.44-5.96) | <0.00* |
| Medications: None | 2.40 (0.58-9.86) | 0.22 |

**OR**, Odds Ratio; **CI**, Confidence Interval.

**Table 5:**
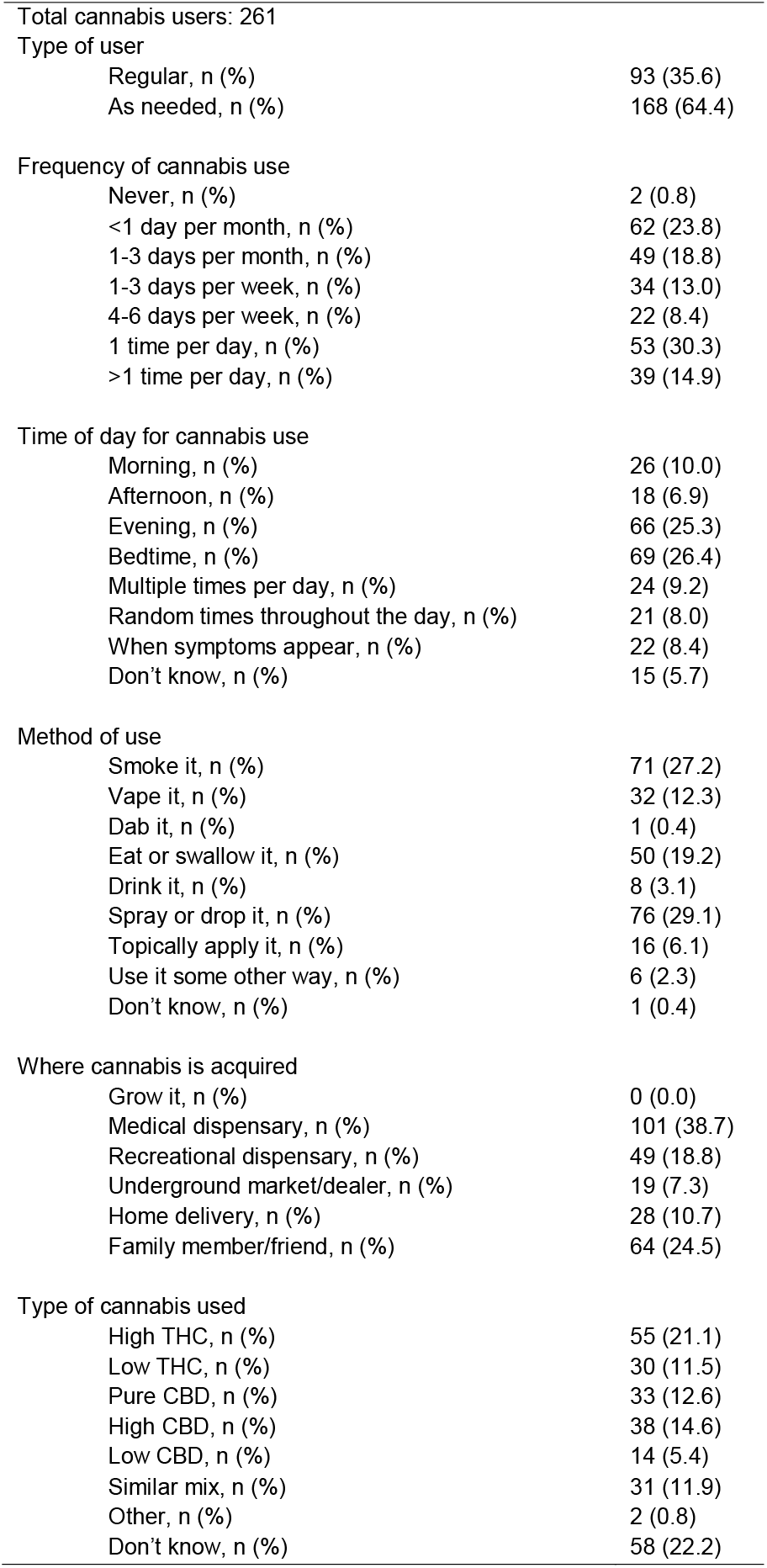
Cannabis Use Methods and Routines

Type of user
|  |  |
| --- | --- |
| Regular, n (%) | 93 (35.6) |
| As needed, n (%) | 168 (64.4) |
| Never, n (%) | 2 (0.8) |
| <1 day per month, n (%) | 62 (23.8) |
| 1-3 days per month, n (%) | 49 (18.8) |
| 1-3 days per week, n (%) | 34 (13.0) |
| 4-6 days per week, n (%) | 22 (8.4) |
| 1 time per day, n (%) | 53 (30.3) |
| >1 time per day, n (%) | 39 (14.9) |
| Time of day for cannabis use |  |
| Morning, n (%) | 26 (10.0) |
| Afternoon, n (%) | 18 (6.9) |
| Evening, n (%) | 66 (25.3) |
| Bedtime, n (%) | 69 (26.4) |
| Multiple times per day, n (%) | 24 (9.2) |
| Random times throughout the day, n (%) | 21 (8.0) |
| When symptoms appear, n (%) | 22 (8.4) |
| Don't know, n (%) | 15 (5.7) |
| Method of use |  |
| Smoke it, n (%) | 71 (27.2) |
| Vape it, n (%) | 32 (12.3) |
| Dab it, n (%) | 1 (0.4) |
| Eat or swallow it, n (%) | 50 (19.2) |
| Drink it, n (%) | 8 (3.1) |
| Spray or drop it, n (%) | 76 (29.1) |
| Topically apply it, n (%) | 16 (6.1) |
| Use it some other way, n (%) | 6 (2.3) |
| Don't know, n (%) | 1 (0.4) |
| Where cannabis is acquired |  |
| Grow it, n (%) | 0 (0.0) |
| Medical dispensary, n (%) | 101 (38.7) |
| Recreational dispensary, n (%) | 49 (18.8) |
| Underground market/dealer, n (%) | 19 (7.3) |
| Home delivery, n (%) | 28 (10.7) |
| Family member/friend, n (%) | 64 (24.5) |
| Type of cannabis used |  |
| High THC, n (%) | 55 (21.1) |
| Low THC, n (%) | 30 (11.5) |
| Pure CBD, n (%) | 33 (12.6) |
| High CBD, n (%) | 38 (14.6) |
| Low CBD, n (%) | 14 (5.4) |
| Similar mix, n (%) | 31 (11.9) |
| Other, n (%) | 2 (0.8) |
| Don't know, n (%) | 58 (22.2) |

**Figure 3:**
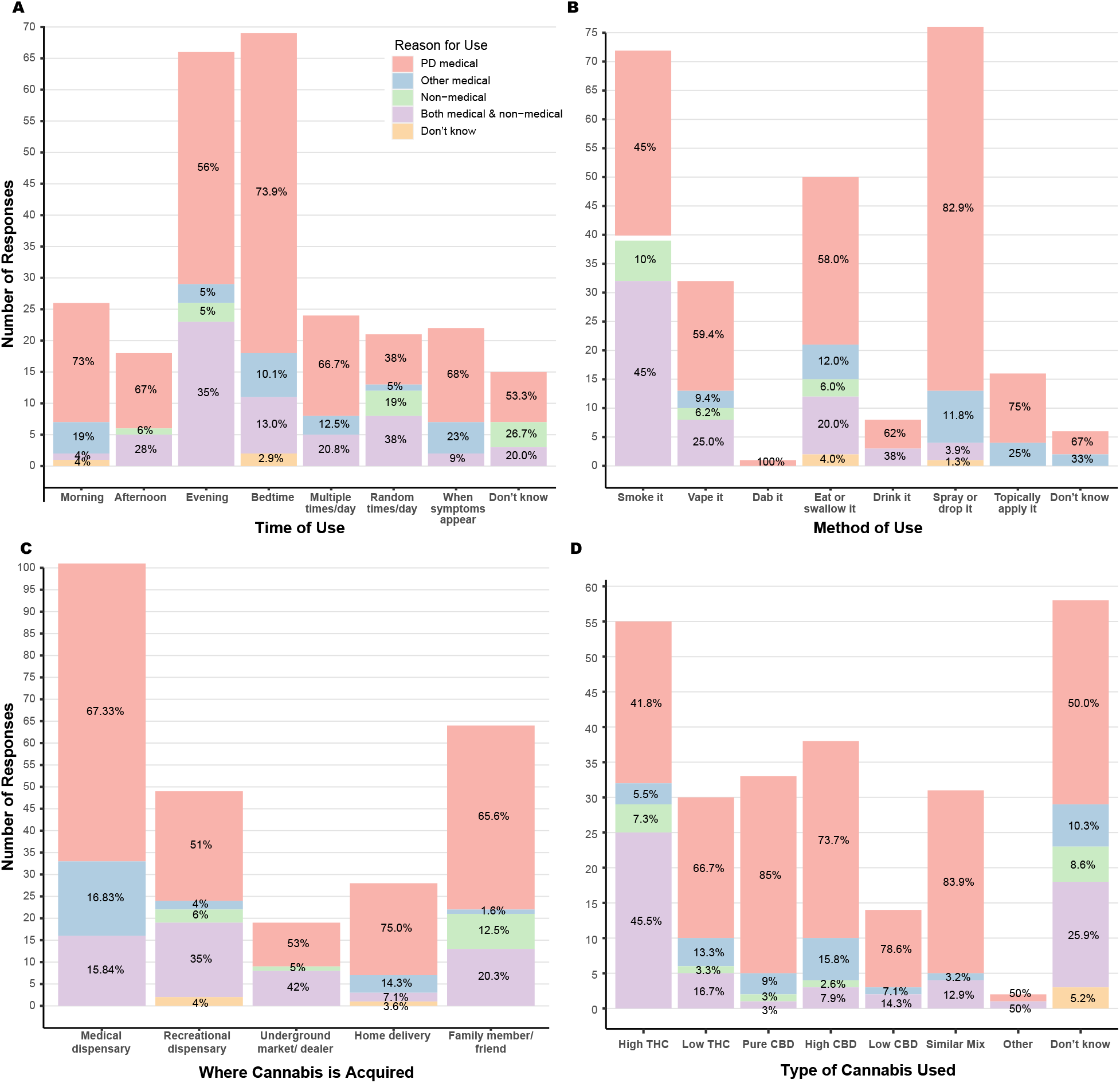
Cannabis use by time of day, method, source, and type. Each cannabis user was asked (**A**) when they used cannabis during the day, (**B**) the primary method by which they used cannabis, (**C**) where they most often acquired their supply of cannabis, and (**D**) the type of cannabis they used based on the concentration of the two main constituents: THC (tetrahydrocannabinol) and CBD (cannabinol). The results were further categorized by type of user (n=261 for all graphs).

Cannabis comes in a number of forms, which can impact how rapidly it is absorbed and for how long it persists in the body.^19^ Users of cannabis most frequently reported spraying or dropping (for example, sublingual drops) (29.1%, 76/261), smoking (27.2%, 71/261) and eating or swallowing (19.2%, 50/261) as their primary method of cannabis use (Fig. 3B). Users acquired cannabis most commonly through a medical dispensary (38.7%, 101/261) or a family member or friend (24.5%, 64/261, see Fig. 3C). When asked details about the type of cannabis used, such as strain of cannabis and concentration of CBD and THC, including high THC, low THC, high CBD, etc., about a quarter of all users did not know (22.2%, 58/261, see Fig. 3D). Of those who did (77.8%, 203/261), almost half did not know the specific type (48.8%, 99/203) or dosage (47.0%, 95/203) they used. Receiving a cannabis recommendation from a licensed provider and identifying as a regular user were associated with knowing the type of cannabis used (OR = 12.2, 95% CI 3.1 to 84.0 and OR = 3.4, 95% CI 1.3 to 12.0 respectively).

### Symptom management by cannabis

We asked all users whether they felt that cannabis addressed their motor and non-motor symptoms. A little less than half reported that its use somewhat addressed their motor (41.0%, 107/261) and non-motor symptoms (42.9%, 112/261, see Table 6). There were no differences for reported efficacy between user groups (PD, other medical, nonmedical, etc.). When differentiated between high THC (n = 51) or high/pure CBD (n = 67) users, high THC users reported better efficacy for both motor and nonmotor symptoms (p = 0.02 and p = 0.001 respectively).

**Table 6:** Reported Symptom Satisfaction Between Cannabis Users and Non-Users, n (%)

|  |  |  |  |
| --- | --- | --- | --- |
| Motor Symptoms | Cannabis among Users<br>(n = 261) | Medications among Users<br>(n = 261) | Medications among Non-users (n = 803) |
| Completely Address | 12 (4.6%) | 9 (3.4%) | 61 (7.6%) |
| Mostly Address | 54 (20.7%) | 126 (48.3%) | 356 (44.3%) |
| Somewhat Address | 107 (41.0%) | 93 (35.6%) | 291 (36.2%) |
| Does Not Address | 88 (33.7%) | 13 (5.0%) | 45 (5.6%) |
| Not Applicable | 0 (0.0%) | 20 (7.7%) | 50 (6.2%) |
| Non-Motor Symptoms |  |  |  |
| Completely Address | 12 (4.6%) | 6 (2.3%) | 51 (6.4%) |
| Mostly Address | 62 (23.8%) | 74 (28.4%) | 276 (34.4%) |
| Somewhat Address | 112 (42.9%) | 122 (46.7%) | 319 (39.7%) |
| Does Not Address | 75 (28.7%) | 43 (16.5%) | 118 (14.7%) |
| Not Applicable | 0 (0.0%) | 16 (6.1%) | 39 (4.9%) |

Respondents who reported using cannabis for general or for specific PD symptoms (90.1%, 200/261) were also asked about perceived symptom improvement. The most common non-motor symptoms that cannabis users were trying to treat were anxiety (45.5%, 91/200), pain (44.0%, 88/200) and sleep disorders (44.0%, 88/200). The most common motor symptoms that cannabis users were trying to treat included stiffness (43.0%, 86/200) and tremor (42.0%, 84/200). Additional symptom information can be found in Fig. 4. When differentiated between high THC or high/pure CBD users, no differences were found between type of cannabis used and the top selected symptoms for anxiety, pain, sleep disturbances, stiffness and tremor. However, there were differences found between type of cannabis used and the selected symptoms, appetite, dystonia and urinary symptoms, but due to the low selection rates, further analysis was not conducted.

**Figure 4:**
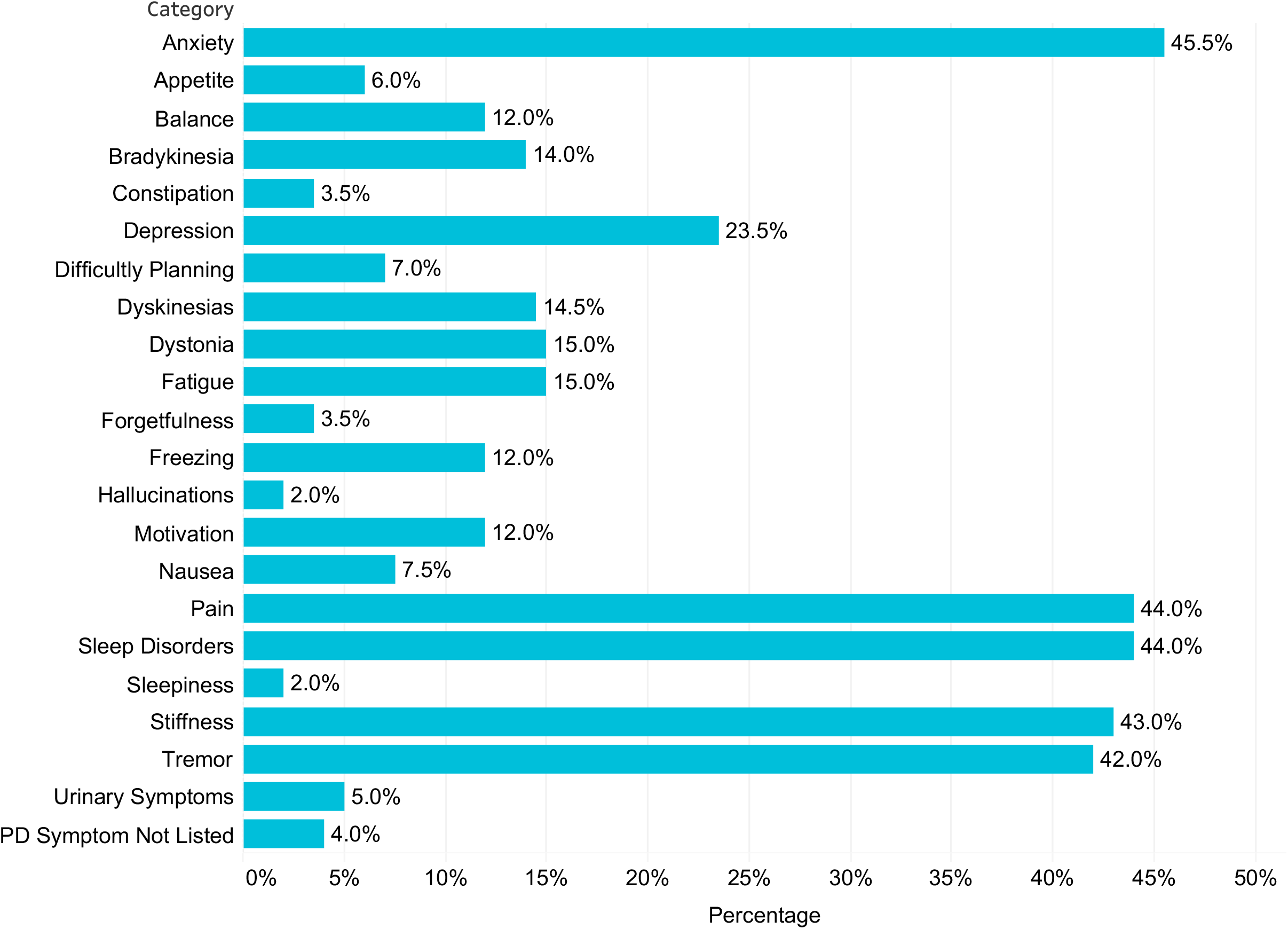
Cannabis use for selected PD symptoms. Users that indicated cannabis use for specific or general PD symptoms (n = 200) were asked, “For which of the following symptoms do you use marijuana or cannabis to improve?”

When asked about associated symptom relief, the majority of these users reported that cannabis use led to a moderate or considerable improvement in the severity of anxiety (78.0%, 71/91), pain (71.6%, 63/88), sleep disorders (76.1%, 67/88), stiffness (64.0%, 55/86) and tremor (63.1%, 53/84). Of those users who selected a slight, moderate or considerable improvement in symptom severity, greater than 80% reported these changes to be meaningful for each of the five symptoms (90.6% anxiety, 89.5% pain, 91.3% sleep disorders, 82.9% stiffness and 84.9% tremor). The majority of these users also reported that cannabis use led to a moderate or considerable improvement in the frequency of anxiety (67.0%, 61/91), pain (63.6%, 56/88), sleep disorders (61.4%, 54/88), stiffness (58.1%, 50/86) and tremor (56.0%, 47/84). Of those users who selected a slight, moderate or considerable improvement in symptom frequency, greater than 80% reported these changes to be meaningful for each of the five symptoms (91.3% anxiety, 87.8% pain, 93.2% sleep disorders, 83.1% stiffness and 93.9% tremor). Reported improvement can be found in Table 7. When differentiated between high THC or high/pure CBD users, there were no significant differences between groups for reported improvement in severity or frequency for sleep, anxiety, stiffness or tremor. However, high THC users did report better improvement for pain severity (p = 0.03) compared to high/pure CBD users.

**Table 7:** Reported Cannabis Symptom Improvement for Top Selected Symptoms, n (%)

|  |  |  |  |  |  |
| --- | --- | --- | --- | --- | --- |
| Improvement to symptom severity | Considerable improvement | Moderate improvement | Slight improvement | No improvement | Worsening of symptoms |
| Severity, n (%) |  |  |  |  |  |
| Anxiety | 32 (35.2) | 39 (42.9) | 14 (15.4) | 5 (5.5) | 1 (1.1) |
| Pain | 31 (35.2) | 32 (36.4) | 13 (14.8) | 12 (13.6) | 0 (0.0) |
| Sleep disorders | 40 (45.5) | 27 (30.7) | 13 (14.8) | 8 (9.1) | 0 (0.0) |
| Stiffness | 22 (25.6) | 33 (38.4) | 21 (24.4) | 8 (9.3) | 2 (2.3) |
| Tremor | 21 (25.0) | 32 (38.1) | 20 (23.8) | 11 (13.1) | 0 (0.0) |
| Was there a meaningful change in QoL? | Yes | No | Don't know |  |  |
| Anxiety | 77 (90.6) | 1 (1.18) | 7 (8.2) |  |  |
| Pain | 68 (89.5) | 3 (4.0) | 5 (6.6) |  |  |
| Sleep disorders | 73 (91.3) | 2 (2.5) | 5 (6.3) |  |  |
| Stiffness | 63 (82.9) | 4 (5.3) | 9 (11.8) |  |  |
| Tremor | 62 (84.9) | 7 (9.6) | 4 (5.5) |  |  |
| Improvement to symptom frequency | Considerable improvement | Moderate improvement | Slight improvement | No improvement | Worsening of symptoms |
| Frequency, n (%) |  |  |  |  |  |
| Anxiety | 29 (31.9) | 32 (35.2) | 19 (20.9) | 10 (11.0) | 1 (1.1) |
| Pain | 28 (31.8) | 28 (31.8) | 18 (20.5) | 14 (15.9) | 0 (0.0) |
| Sleep disorders | 37 (42.0) | 17 (19.3) | 20 (22.7) | 14 (15.9) | 0 (0.0) |
| Stiffness | 20 (23.3) | 30 (34.9) | 21 (24.4) | 14 (16.3) | 1 (1.2) |
| Tremor | 23 (27.4) | 24 (28.6) | 19 (22.6) | 18 (21.4) | 0 (0.0) |
| Was there a meaningful change in QoL? | Yes | No | Don't know |  |  |
| Anxiety | 73 (91.3) | 1 (1.3) | 7 (7.5) |  |  |
| Pain | 65 (87.8) | 5 (6.8) | 4 (5.4) |  |  |
| Sleep disorders | 69 (93.2) | 1 (1.4) | 4 (5.4) |  |  |
| Stiffness | 59 (83.1) | 3 (4.2) | 9 (12.7) |  |  |
| Tremor | 62 (93.9) | 2 (3.0) | 2 (3.0) |  |  |

### Cannabis user experiences

A small portion of users (12.6% 33/261) reported negative side effects from cannabis use. When differentiated between high THC and high/pure CBD, high THC users were 7.43 times more likely to report side effects than high/pure CBD users (95% CI 1.38 to 59.76). We also polled non-users of cannabis who had indicated a previous negative experience as a reason for not using cannabis in the past six months (4.1%, 33/803). Of these non-users, about half (51.5%, 17/33) also reported negative side effects from cannabis use. These two combined sub-groups most commonly identified anxiety (30.0%, 15/50), impaired coordination (20.0%, 10/50), dizziness (20.0%, 10/50), and “other reasons” (38.0%, 19/50; open text responses included sleepiness, confusion, worsening orthostatic hypotension, etc.) as side effects of cannabis use.

About a quarter of cannabis users (23.0%, 60/261) had stopped using cannabis in the previous six months. Among these users as well as the subset of non-users who had indicated a previous negative experience (4.1%, 33/803), a lack of symptom improvement (35.5%, 33/93) was the most commonly selected reason for discontinued use (Fig. 5). Identifying as a regular user was inversely associated with cessation (OR = 0.1, 95% CI 0.01 to 0.6). Although high THC users were more likely to report side effects, there were no differences in cessation rates between high THC and high/pure CBD users. Among users who had not discontinued use in the previous six months, the most commonly identified items that cannabis use allowed for included general symptom improvement (40.3%, 81/201), relaxation (14.4%, 29/201) and improvement in daily activities (9.0%, 18/201).

**Figure 5:**
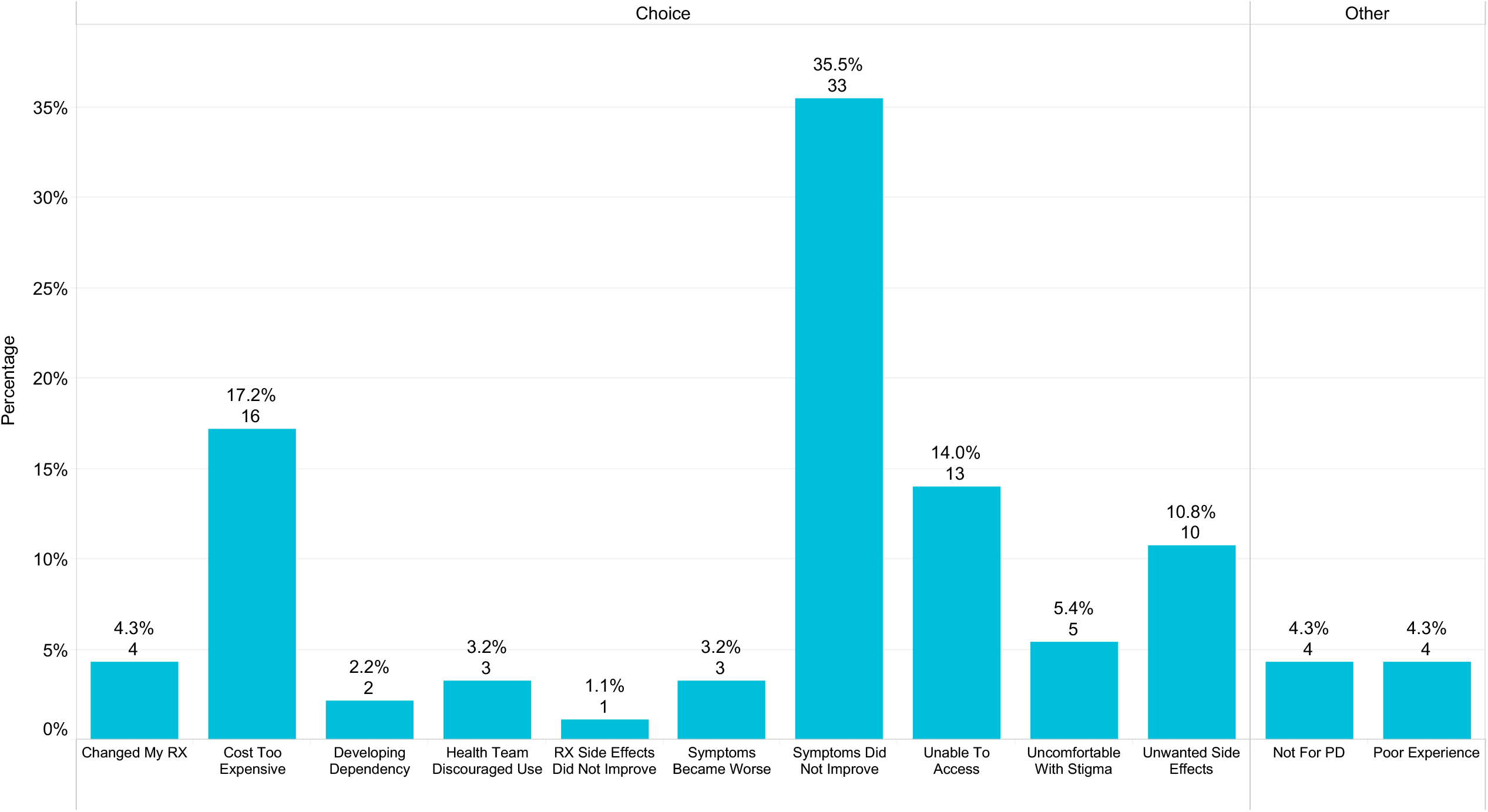
Reported reason for cannabis cessation. Users and former users (n = 93) responded to the survey question “For what reason did you stop using marijuana or cannabis?” Responses were either pre-identified as one of the survey options or individuals selected “other” and their open text response was analyzed thematically. Lack of symptom improvement was a key reason for halting cannabis use.

### Medication satisfaction

All respondents (both users and non-users) reported on their prescription medication use. A few reported not taking medications for PD motor (6.6%, 70/1064) or for non-motor (5.2%, 55/1064) symptoms. Very few (4.6%, 49/1064) were taking no medication at all. Longer disease duration and being male were both inversely associated with being medication free (OR = 0.7, 95% CI 0.5 to 0.9 and OR = 0.1, 95% CI 0.01 to 0.5 respectively). However, those not taking PD prescription medications were no more or less likely to report cannabis usage.

We asked those taking PD medications whether their prescription medications addressed their motor and non-motor symptoms. For motor symptom management, about half (45.3%, 482/1064) reported that medications mostly addressed their symptoms. For non-motor symptom management, a little less than half (41.5%, 441/1064) reported that medications somewhat addressed their symptoms (Table 6). There were no reported differences between users and non-users regarding reported satisfaction for motor symptoms; however, there were significant differences between users and non-users for non-motor symptoms (p = 0.01). Users rated their prescription medications with higher efficacy than cannabis for control of both motor and non-motor symptoms (p≪0.001 and p = 0.03 respectively).

Among all users, the majority (84.7%, 221/261) reported that cannabis use had no impact on their prescription medication usage, and of the users who had indicated cannabis use for both medical reasons and medical and non-medical reasons, most (89.0%, 219/246) reported that they had not thought that cannabis would be a replacement for their PD prescription medications when they began using.

### Clinical Trials

We queried all respondents (both users and non-users) about their level of interest in learning about and enrolling in a clinical trial exploring the impact of cannabis on PD symptoms. Most (82.3%, 876/1064) were interested in learning more information about a clinical trial, and more than half (62.3%, 663/1064) were interested in enrolling in a clinical trial. Upon further analysis, interest in clinical trial enrollment was associated with cannabis use, and a longer disease duration (OR = 3.2, 95% CI 2.2 to 4.8 and OR = 1.03, 95% CI 1.01 to 1.06 respectively).

## Discussion

We examined cannabis use, knowledge, motivations, and routines among people living with PD. This study is unique in that we captured both medicinal and recreational cannabis use among a large number of people living with PD from nearly all fifty states. From our population sample, nearly a quarter of those surveyed (24.5%) reported the use of cannabis. This is within the wide range of cannabis usage reported among people living with PD in the literature (between 4% and 80%).^10,11,14,20,21^ However, our reported number of users may be higher because we assessed both the recreational and medicinal use of cannabis while other studies cited assessed medicinal use only.

Our results suggest that although there are many people with PD using cannabis as a CAIM treatment for their motor and non-motor symptoms, the lack of formal guidance about cannabis usage for PD may underlie inconsistencies in use and reported effectiveness. The survey results also indicate a knowledge gap among people living with PD, which may be impacting their decisions about cannabis use. In particular, 22.2% of users did not know the type or form of cannabis; 47% did not know their dosage, and, of those that reported a type of cannabis (high THC, pure CBD, etc.), 48.8% did not know the name of what they used. These findings are similar to a recent study reporting that 17.7% of respondents had not known that type of cannabis used.^21^

This knowledge gap, combined with the receipt of information and recommendations primarily through the internet, friends or other people living with PD or word of mouth, as opposed to a licensed health provider, could be partially responsible for the perceived confusion about expectations and efficacy, as not knowing the type of cannabis used and cessation were both associated with provider recommendations. We would expect that specific cannabis formulations, ratio of CBD to THC, route of administration, and strain (*sativa* vs *indica*) might have a differential impact of specific symptoms that patients are trying to address.^12,13,19^ With these factors ignored or unknown and without an individualized and educated approach, the potential for success may be limited and the potential for adverse reactions may be amplified. However, in this study, users who did not know the type of cannabis they used compared to those who did know did not report significantly different levels to which cannabis addressed motor and non-motor symptoms.

Among some users, reported cannabis use may have been beneficial for particular motor and non-motor PD symptoms including anxiety, pain, sleep disorders, stiffness and tremor. These symptoms are consistent with other research studies examining motor and non-motor effects of cannabis on PD symptoms.^11,14,21–26^ In this study, users of cannabis reported it most improved the severity of anxiety, pain and sleep disorders. However, overall use and the level to which cannabis was reported to address motor and non-motor symptoms varied widely. This was particularly evident by both the reported low levels of motor and non-motor symptom relief by cannabis and the 23.0% discontinuance rate in the previous six months, which was greater than the 14% reported in a survey of MS patients.^27^ Here, discontinued use was primarily reported due to a lack of symptom improvement. These findings are in line with the inconsistent efficacy observed in the limited clinical trials conducted on cannabis and PD.^12,28,29^

We found that a majority of people living with PD and using cannabis, were doing so without having been provided information or provider recommendations. This was not entirely surprising; a previous study found a significant cannabis knowledge gap among specialized Parkinson’s disease clinicians that paralleled our observed knowledge gap in people with PD. In that survey; the majority of clinicians reported they lacked the formal training and knowledge of cannabis as a treatment alternative for PD motor and non-motor symptoms.^14^

It is important that people living with PD are provided information from reputable, evidence-based sources. Considering the growing legality of cannabis access and customization of cannabis formulations marketed to people living with PD, additional research is needed for both clinicians and people with PD to better understand the efficacy of cannabis for PD motor and non-motor symptoms. If cannabis is intended to be used as a medicinal or CAIM treatment for PD, formal recommendations are needed to fill the current knowledge gap around cannabis type, dosage and efficacy. Any future research or clinical trials targeting the efficacy of cannabis should clearly define these recommendations for use and prioritize the communication of such recommendations to enrolled participants.

The large number of non-users reported here suggest that cannabis use may not be viewed by people living with PD as a primary medicinal treatment option for PD. Although 33 states have legalized cannabis use for medical conditions, PD is recognized as a qualifying medical condition in only 17 of these states.^12^ In this study, 36.4% of cannabis users had other medical, recreational or a combination of reasons for cannabis use, and the identified reasons for use were associated with variability in responses to use routines.

Cannabis use did not have an impact on PD prescription medications. Indeed, nearly all respondents (89%) reported that they knew cannabis would not be a replacement for their current medications. However this finding differs from other studies, in which prescription medication use decreased among cannabis users.^20^ Despite the perceived hype around cannabis for PD,^18^ a healthy skepticism seems apparent among non-users.^21^ Nearly three-quarters (76%) would consider cannabis use only if there were scientific evidence to support the claimed benefits. Our results would suggest that people living with PD expect and could benefit from more evidence about use and outcomes.

It is notable that pain is one of the symptoms best described in the literature that has evidence supporting use of cannabis in other populations such as HIV neuropathy and cancer related pain.^30,31^ However, those results have been recently called into question.^32^ Supporting the point that patients look towards evidence, here, one of the main uses reported for cannabis among those with PD was for the amelioration of pain symptoms. As evidence is generated around the efficacy of cannabis for PD, such evidence is expected to influence how cannabis use is viewed as a PD treatment option in the future.

There are several limitations to this study. Respondents were acquired through a convenience sample, and as a result the demographics reflected in this study primarily consisted of white, married and highly educated respondents, which likely do not reflect the entire PD population. The study response rate of 17.6% also suggests that respondents may not have been representative of the PD population. Another limitation is that cannabis users may have been more inclined to respond to the survey and report favorable results than non-users, which could have influenced and inflated the number of users and benefits leading to a possible response bias. However, the large number of non-users also participating may have mitigated this potential response bias by users. All respondents were required to complete the survey electronically, and this survey invitation was sent only to those who had an active email address.

Given that PD predominantly affects the older population, it is possible that the technology and email requirements excluded several who did not have an active email address or access to an electronic or mobile device. It is similarly noteworthy that the average educational level (and presumed socioeconomic status) of the respondents was high. Thus, our respondents may be more motivated to participate in such surveys, more comfortable with the use of electronic technology, and potentially less concerned about legal ramifications of their participation. Although cannabis is recreationally legal in 11 states and medicinally legal in 33 states,^12^ it remains a United States Drug Enforcement Administration Schedule I substance. For this reason, people living with PD may have been reluctant to report their cannabis use, leading to an undercount of user prevalence. Policy around cannabis varies globally and use likely does so as well. This study was limited to the United States and is therefore not reflective of other countries’ cannabis use trends or perceptions.

In summary, a large survey of individuals with PD regarding their attitudes and knowledge of cannabis revealed that 1 in 5 of people with PD are current cannabis users. Reassuringly, most of these users recognize that cannabis is not a substitute for their current medications and also see the limited efficacy of cannabis for symptom management. Many people with PD justifiably are not using cannabis because of a lack of evidence to support its use with PD. Despite the lack of evidentiary support, it is a concern that PD cannabis users lack resources to guide them on the potential use of cannabis for PD. PD cannabis users identify several non-motor symptoms that may be responsive to cannabis and should be the target of future clinical experiments to see if this reported experience bares out in a larger population.

## Methods

### Standard Protocol Approvals, Registrations, and Patient Consents

The Western Institutional Review Board (WIRB) approved this study. Given the low-risk nature of the protocol and anonymous data collection, the IRB found that this research meets the requirements for a waiver of consent under 45 CFR 46 116 (f) [2018 Requirements] 45 CRF 46.116(d) [Pre-2018 Requirements].

### Patient Surveys

A review of publicly available surveys about cannabis use or perceptions of use was conducted, and relevant questions were used or modified to fit this survey.^14,33–37^ The survey was then reviewed by three people living with PD to ensure relevance and readability.

An invitation to an anonymous, electronic survey was emailed on January 17, 2020 to 7,607 people with Parkinson’s disease. These individuals were identified as having attended an online or in-person Parkinson’s Foundation educational event or having called the Parkinson’s Foundation Helpline in the 2019 calendar year. Reminder emails, two in total, were sent a week after the initial invitation and a week before the survey closed. The survey remained open for three weeks and closed on February 7, 2020.

The online questionnaire consisted of seven sections:

1. A screening question to confirm that the respondent was a person with Parkinson’s disease *(number of questions, n = 1)*.
2. A question about cannabis usage in the last six months. Due to possible confusion about the definition of cannabis, both the terms marijuana and cannabis were included in all questions throughout the survey *(n =1)*.
3. Questions about reasons for not using cannabis *(n =2)*.
4. Questions about motivations for cannabis use, information sources use, frequency and methods of use, perceived PD symptom or medication side effect improvement, perceived cannabis side effects, cessation of cannabis use and motivations for cessation *(n =16)*.
5. Questions about PD prescription medications and perceived impact and interactions on PD prescription medications *(n =4)*.
6. Questions about interest in future cannabis clinical trials *(n =2)*.
7. Basic demographic information including year of birth and diagnosis, marital status, gender, employment status, etc. *(n =11)*.

The full questionnaire can be viewed in the Supplementary Information.

Of the 1,339 surveys returned, 261 were incomplete and were discarded from further analysis. There were 1,078 completed responses; however, five were discarded due to incomplete or incorrect geographic information (e.g. one, four or eight digit zip codes) and an additional nine were discarded due to suspected inaccuracy of birth year and diagnosis year (e.g. reported year of birth after 2010 or reported year of diagnosis equal to reported year of birth). 1,064 completed responses were available for analysis. However, when results were further analyzed by gender, five respondents who had selected other or prefer not to answer as their gender were removed from the gender analysis portion of the reported results.

### Statistical Analysis

The anonymous survey was collected and managed using Research Electronic Data Capture (REDCap) tools hosted at the Parkinson’s Foundation.^38,39^ Statistical analyses were performed using the R software package and programming language through RStudio desktop version 1.2.5042.^40,41^ Determination of associations between demographics, user status and reported outcomes were performed using Student’s t-test for continuous variables, Pearson’s Chi-squared test (including the Monte Carlo procedure for p-value approximation where cell counts were below 5) and Kruskal-Wallis test for categorical variables. Variables rating demographic characteristics, user stats and reported outcomes that were dichotomous or could be dichotomized were examined using Mann-Whitney U test and multivariate logistic regression. *P* < 0.05 was considered statistically significant. All tests were two-sided. Open text collected in the survey were evaluated through thematic analysis.

## Data Availability

The datasets generated during and/or analyzed during the current study are available from the corresponding author on reasonable request.

## Code Availability

Statistical analysis was performed using the R software package and programming language through RStudio desktop version 1.2.5042. R syntax used for the analysis is available from the corresponding author upon reasonable request.

## Acknowledgements

This study was supported by the generous donors of the Parkinson’s Foundation. The authors would like to acknowledge Parkinson’s Foundation Research Advocates Steven Gilbert, Andre Hosang and Elizabeth Ogren for their contributions to survey review from their perspectives of people living with Parkinson’s disease. We would also like to thank Tom Borger and his team at Ilera Healthcare for sharing sample cannabis-related survey questions.

## Author Contributions

M.P.F.: research project organization and execution; survey design; execution and review of statistical analysis; and writing of first draft of the manuscript. D.B., B.M.K., A.J.S., and C.M.E.: survey design and review of manuscript. R.D.L: IRB protocol execution and review of the manuscript. J.C.B.: research project conception; survey design; review of statistical analysis; and review of the manuscript.

## Competing Interests

The authors declare no competing interests.

## Notes

### Competing Interest Statement

The authors have declared no competing interest.

### Funding Statement

The authors received no specific funding for this work.

### Author Declarations

The Western Institutional Review Board (WIRB) approved this study. Given the low-risk nature of the protocol and anonymous data collection, the Board found that this research meets the requirements for a waiver of consent under 45 CFR 46 116 (f) [2018 Requirements] 45 CRF 46.116(d) [Pre-2018 Requirements].

