## Supplementary Table for "Weeding Through the Haze: A Survey on Cannabis Use Among People Living with Parkinson’s Disease in the US"

Supplementary Information: Cannabis Survey

| Variable / Field Name | Section Header | Field Type | Field Label | Choices, Calculations, OR Slider Labels | Branching Logic (Show field only if...) | Required Field? |
|---|---|---|---|---|---|---|
| participant_id | | text | Participant ID | | | |
| pwp | | radio | Are you a person with Parkinson's disease? | 1, Yes \| 0, No | | y |
| redirect | | descriptive | Thank you for sharing your feedback. Unfortunately, this survey is only intended for people with Parkinson’s to complete. Please reach out to Senior Manager, Community Engagement, Megan Feeney, MPH at 646-388-7674 or with any questions. | | [pwp] = '0' | |
| one | Marijuana or Cannabis Usage | radio | Have you used marijuana or cannabis at least once in the past six months? | 1, Yes \| 0, No | [pwp] = '1' | y |
| one_a | | checkbox | What were your primary reasons for not using marijuana or cannabis in the past six months? Select up to three. | 1, Fear of potential side effects or adverse outcomes from using marijuana or cannabis \| 2, Cost of marijuana or cannabis \| 3, Lack of scientific evidence about benefits of using marijuana or cannabis \| 4, Stigma associated using marijuana or cannabis \| 5, Negative previous experience using marijuana or cannabis \| 6, Use discouraged by my health team \| 7, Other | [pwp] = '1' and [one] = '0' | y |
| one_a_a | | notes | What were your other reasons for not using marijuana or cannabis in the past six months? | | [one_a(7)] = '1' | y |
| one_b | | checkbox | For which of the following reasons would you consider using marijuana or cannabis? Select up to three of the most influential reasons. | 1, Potential side effects and adverse outcomes of using marijuana or cannabis were better understood \| 2, The cost of marijuana or cannabis was partially or fully covered by health insurance \| 3, Scientific evidence supported the claimed benefits of using marijuana or cannabis \| 4, There was less stigma associated with using marijuana or cannabis \| 5, Your health team encouraged you to try marijuana or cannabis \| 6, Marijuana or cannabis was legalized in your state \| 7, Other | [pwp] = '1' and [one] = '0' | y |
| one_b_a | | notes | For what other reasons would you consider using marijuana or cannabis? | | [one_b(7)] = '1' | y |
| two | | radio | When you used marijuana or cannabis during the past six months, was it usually: | 1, For PD related medical reasons (for example, to treat or decrease symptoms of PD) \| 2, For other medical reasons (for example, to treat or decrease symptoms of another health condition) \| 3, For nonmedical reasons (for example, recreational use) \| 4, For both medical and nonmedical reasons \| 5, Don't know/Not sure | [pwp] = '1' and [one] = '1' | y |
| two_a | | radio | Where did you learn about marijuana or cannabis use for these reasons? | 1, Doctor \| 2, Internet/News \| 3, Media/advertisements \| 4, PD foundation \| 5, PD support groups \| 6, Friends or other people with PD \| 7, Family \| 8, Other | [two] = '1' or [two] = '2' or [two] = '4' | y |

| | | | | | | |
|---|---|---|---|---|---|---|
| two_a_a | | text | Where else did you learn about marijuana or cannabis use for these reasons? | | [two_a] = '8' | y |
| two_b | | radio | Was marijuana or cannabis recommended to you by a licensed doctor or provider? | 1, Yes \| 0, No | [two] = '1' or [two] = '2' or [two] = '4' | y |
| two_c | | radio | Were you provided information about recommendations for marijuana or cannabis use (for example, dosage, type, frequency of use, where it can be accessed, etc.)? | 1, Yes \| 0, No | [two] = '1' or [two] = '2' or [two] = '4' | y |
| two_c_c | | radio | From whom were you provided information about recommendations for marijuana or cannabis use? | 1, My PD doctor \| 2, Another licensed doctor (for example, primary care physician) \| 3, Staff at a medical dispensary \| 4, Friends or other people with PD \| 5, Family \| 6, Other | [two_c] = '1' | y |
| two_c_c_c | | text | From who else were you provided information about recommendations for marijuana or cannabis use? | | [two_c_c] = '6' | y |
| two_d | | radio | What is your primary reason for using marijuana or cannabis for PD? | 1, To improve specific PD symptoms \| 2, To improve PD symptoms in general \| 3, To decrease PD medication (i.e. Sinemet, Mirapex) side effects \| 4, I have no primary reason for using marijuana or cannabis for PD medical reasons | [two] = '1' or [two] = '4' | y |
| two_e | | text | For what medical condition do you primarily use marijuana or cannabis? | | [two] = '2' | y |
| three | | radio | What kind of marijuana or cannabis user would you consider yourself to be? | 1, Regular user \| 2, Occasional or "as needed" user | [pwp] = '1' and [one] = '1' | y |
| four | | radio | In the past six months, which of the following best captures your average frequency of marijuana or cannabis use? | 1, Never \| 2, Less than one day per month \| 3, One to three days per month \| 4, One to three days per week \| 5, Four to six days per week \| 6, Once per day \| 7, Multiple times per day | [pwp] = '1' and [one] = '1' | y |
| five | | radio | Is there a particular time of day you most often use marijuana or cannabis? | 1, Morning \| 2, Afternoon \| 3, Evening \| 4, Bedtime \| 5, I use marijuana or cannabis multiple times per day \| 6, I use marijuana or cannabis at random times throughout the day \| 7, I use marijuana when specific symptoms appear \| 8, Don't know/Not sure | [pwp] = '1' and [one] = '1' | y |
| six | | radio | During the past six months, which one of the following ways did you use marijuana or cannabis the most often? Did you usually... | 1, Smoke it \| 2, Vaporize it \| 3, Dab it (for example, using waxes or concentrates) \| 4, Eat or swallow it (for example, edibles or capsules) \| 5, Drink it (for example, in tea) \| 6, Spray or drop it under tongue (for example, sublingual drops or dissolvable strips) \| 7, Topically apply it (for example, creams or lotions) \| 8, Wear it (for example, patch) \| 9, Use it some other way \| 10, Don't know/not sure | [pwp] = '1' and [one] = '1' | y |

| | | | | | | |
|---|---|---|---|---|---|---|
| six_a | | text | What other way did you use marijuana or cannabis the most often? | | [six] = '9' | y |
| seven | | radio | Where do you most often get your marijuana or cannabis from? | 1, Grow it myself \| 2, Medical dispensary \| 3, Recreational dispensary \| 4, Underground market/ dealer \| 5, Home delivery \| 6, Family member/Friend | [pwp] = '1' and [one] = '1' | y |
| eight | | radio | What type of marijuana or cannabis do you most often use? | 1, High THC content \| 2, Low THC content \| 3, Pure CBD \| 4, High CBD content \| 5, Low CBD content \| 6, Similar or equal mix of THC and CBD \| 7, Other \| 8, Don't know | [pwp] = '1' and [one] = '1' | y |
| eight_z | | text | What other type of marijuana or cannabis do you most often use? | | [eight] = '7' | |
| eight_a | | text | What is the specific type of marijuana or cannabis you most often use (for example, street name or brand name)? If you do not know, please write "Don't know." | | [eight] = '1' or [eight] = '2' or [eight] = '3' or [eight] = '4' or [eight] = '5' or [eight] = '6' or [eight] = '7' | y |
| eight_b | | text | What is the specific dosage or potency of marijuana or cannabis you most often use? If you do not know, please write "Don't know." | | [eight] = '1' or [eight] = '2' or [eight] = '3' or [eight] = '4' or [eight] = '5' or [eight] = '6' or [eight] = '7' | y |
| nine | Intended Symptoms and Side Effects to Treat and Associated Outcomes | checkbox | For which of the following symptoms do you use marijuana or cannabis to improve? Select all that apply. | 1, Appetite \| 2, Pain \| 3, Nausea \| 4, Anxiety \| 5, Tremor \| 6, Stiffness \| 7, Dyskinesias \| 8, Depression \| 9, Fatigue \| 10, Freezing \| 11, Constipation \| 12, Motivation \| 13, Sleepiness (daytime) \| 14, Sleep disorders \| 15, Dystonia \| 16, Bradykinesia (slowness of movement) \| 17, Urinary symptoms \| 18, Balance \| 19, Forgetfulness \| 20, Difficulty planning and/or multi-tasking \| 21, Hallucinations \| 22, My PD symptom is not listed on this list. \| 23, I do not use marijuana or cannabis for specific PD symptoms | [two_d] = '2' or [two_d] = '1' | y |
| nine_a | | text | For what other symptom do you use marijuana or cannabis to improve? | | [nine(22)] = '1' | y |
| nine_9_appetite | | radio | To what extent do you feel that marijuana or cannabis improved the severity of appetite loss? | 1, Considerable improvement \| 2, Moderate improvement \| 3, Slight improvement \| 4, No improvement \| 5, Worsening of symptoms | [nine(1)] = '1' | y |
| nine_a_a_appetite | | radio | Were these changes meaningful to your quality of life? | 1, Yes \| 0, No \| 2, Don't know | [nine_9_appetite] = '2' or [nine_9_appetite] = '1' or [nine_9_appetite] = '3' | y |
| nine_9_pai n | | radio | To what extent do you feel that marijuana or cannabis improved the severity of pain? | 1, Considerable improvement \| 2, Moderate improvement \| 3, Slight improvement \| 4, No improvement \| 5, Worsening of symptoms | [nine(2)] = '1' | y |
| nine_a_a_pain | | radio | Were these changes meaningful to your quality of life? | 1, Yes \| 0, No \| 2, Don't know | [nine_9_pain] = '2' or [nine_9_pain] = '1' or [nine_9_pain] = '3' | y |
| nine_9_nausea | | radio | To what extent do you feel that marijuana or cannabis improved the severity of nausea? | 1, Considerable improvement \| 2, Moderate improvement \| 3, Slight improvement \| 4, No improvement \| 5, Worsening of symptoms | [nine(3)] = '1' | y |

| | | | | | |
|---|---|---|---|---|---|
| nine_a_a_nausea | radio | Were these changes meaningful to your quality of life? | 1, Yes \| 0, No \| 2, Don't know | [nine_9_nausea] = '2' or [nine_9_nausea] = '1' or [nine_9_nausea] = '3' | y |
| nine_9_anxiety | radio | To what extent do you feel that marijuana or cannabis improved the severity of anxiety? | 1, Considerable improvement \| 2, Moderate improvement \| 3, Slight improvement \| 4, No improvement \| 5, Worsening of symptoms | [nine(4)] = '1' | y |
| nine_a_a_anxiety | radio | Were these changes meaningful to your quality of life? | 1, Yes \| 0, No \| 2, Don't know | [nine_9_anxiety] = '2' or [nine_9_anxiety] = '1' or [nine_9_anxiety] = '3' | y |
| nine_9_tremor | radio | To what extent do you feel that marijuana or cannabis improved the severity of tremor? | 1, Considerable improvement \| 2, Moderate improvement \| 3, Slight improvement \| 4, No improvement \| 5, Worsening of symptoms | [nine(5)] = '1' | y |
| nine_a_a_tremor | radio | Were these changes meaningful to your quality of life? | 1, Yes \| 0, No \| 2, Don't know | [nine_9_tremor] = '2' or [nine_9_tremor] = '1' or [nine_9_tremor] = '3' | y |
| nine_9_stiffness | radio | To what extent do you feel that marijuana or cannabis improved the severity of stiffness? | 1, Considerable improvement \| 2, Moderate improvement \| 3, Slight improvement \| 4, No improvement \| 5, Worsening of symptoms | [nine(6)] = '1' | y |
| nine_a_a_stiffness | radio | Were these changes meaningful to your quality of life? | 1, Yes \| 0, No \| 2, Don't know | [nine_9_stiffness] = '2' or [nine_9_stiffness] = '1' or [nine_9_stiffness] = '3' | y |
| nine_9_dyskinesias | radio | To what extent do you feel that marijuana or cannabis improved the severity of dyskinesias? | 1, Considerable improvement \| 2, Moderate improvement \| 3, Slight improvement \| 4, No improvement \| 5, Worsening of symptoms | [nine(7)] = '1' | y |
| nine_a_a_dyskinesias | radio | Were these changes meaningful to your quality of life? | 1, Yes \| 0, No \| 2, Don't know | [nine_9_dyskinesias] = '2' or [nine_9_dyskinesias] = '1' or [nine_9_dyskinesias] = '3' | y |
| nine_9_depression | radio | To what extent do you feel that marijuana or cannabis improved the severity of depression? | 1, Considerable improvement \| 2, Moderate improvement \| 3, Slight improvement \| 4, No improvement \| 5, Worsening of symptoms | [nine(8)] = '1' | y |
| nine_a_a_depression | radio | Were these changes meaningful to your quality of life? | 1, Yes \| 0, No \| 2, Don't know | [nine_9_depression] = '2' or [nine_9_depression] = '1' or [nine_9_depression] = '3' | y |
| nine_9_fatigue | radio | To what extent do you feel that marijuana or cannabis improved the severity of fatigue? | 1, Considerable improvement \| 2, Moderate improvement \| 3, Slight improvement \| 4, No improvement \| 5, Worsening of symptoms | [nine(9)] = '1' | y |
| nine_a_a_fatigue | radio | Were these changes meaningful to your quality of life? | 1, Yes \| 0, No \| 2, Don't know | [nine_9_fatigue] = '2' or [nine_9_fatigue] = '1' or [nine_9_fatigue] = '3' | y |

| | | | | | |
|---|---|---|---|---|---|
| nine_9_freezing | radio | To what extent do you feel that marijuana or cannabis improved the severity of freezing? | 1, Considerable improvement \| 2, Moderate improvement \| 3, Slight improvement \| 4, No improvement \| 5, Worsening of symptoms | [nine(10)] = '1' | y |
| nine_a_a_freezing | radio | Were these changes meaningful to your quality of life? | 1, Yes \| 0, No \| 2, Don't know | [nine_9_freezing] = '2' or [nine_9_freezing] = '1' or [nine_9_freezing] = '3' | y |
| nine_9_constipation | radio | To what extent do you feel that marijuana or cannabis improved the severity of constipation? | 1, Considerable improvement \| 2, Moderate improvement \| 3, Slight improvement \| 4, No improvement \| 5, Worsening of symptoms | [nine(11)] = '1' | y |
| nine_a_a_constipation | radio | Were these changes meaningful to your quality of life? | 1, Yes \| 0, No \| 2, Don't know | [nine_9_constipation] = '2' or [nine_9_constipation] = '1' or [nine_9_constipation] = '3' | y |
| nine_9_motivation | radio | To what extent do you feel that marijuana or cannabis improved the severity of motivation loss? | 1, Considerable improvement \| 2, Moderate improvement \| 3, Slight improvement \| 4, No improvement \| 5, Worsening of symptoms | [nine(12)] = '1' | y |
| nine_a_a_motivation | radio | Were these changes meaningful to your quality of life? | 1, Yes \| 0, No \| 2, Don't know | [nine_9_motivation] = '2' or [nine_9_motivation] = '1' or [nine_9_motivation] = '3' | y |
| nine_9_sleepiness_daytime | radio | To what extent do you feel that marijuana or cannabis improved the severity of sleepiness (daytime)? | 1, Considerable improvement \| 2, Moderate improvement \| 3, Slight improvement \| 4, No improvement \| 5, Worsening of symptoms | [nine(13)] = '1' | y |
| nine_a_a_sleepiness_daytime | radio | Were these changes meaningful to your quality of life? | 1, Yes \| 0, No \| 2, Don't know | [nine_9_sleepiness_daytime] = '2' or [nine_9_sleepiness_daytime] = '1' or [nine_9_sleepiness_daytime] = '3' | y |
| nine_9_sleepdisorders | radio | To what extent do you feel that marijuana or cannabis improved the severity of sleep disorders? | 1, Considerable improvement \| 2, Moderate improvement \| 3, Slight improvement \| 4, No improvement \| 5, Worsening of symptoms | [nine(14)] = '1' | y |
| nine_a_a_sleepdisorders | radio | Were these changes meaningful to your quality of life? | 1, Yes \| 0, No \| 2, Don't know | [nine_9_sleepdisorders] = '2' or [nine_9_sleepdisorders] = '1' or [nine_9_sleepdisorders] = '3' | y |
| nine_9_dystonia | radio | To what extent do you feel that marijuana or cannabis improved the severity of dystonia? | 1, Considerable improvement \| 2, Moderate improvement \| 3, Slight improvement \| 4, No improvement \| 5, Worsening of symptoms | [nine(15)] = '1' | y |

| | | | | | |
|---|---|---|---|---|---|
| nine_a_a_dystonia | radio | Were these changes meaningful to your quality of life? | 1, Yes \| 0, No \| 2, Don't know | [nine_9_dystonia] = '2' or [nine_9_dystonia] = '1' or [nine_9_dystonia] = '3' | y |
| nine_9_bradykinesia | radio | To what extent do you feel that marijuana or cannabis improved the severity of bradykinesia (slowness of movement)? | 1, Considerable improvement \| 2, Moderate improvement \| 3, Slight improvement \| 4, No improvement \| 5, Worsening of symptoms | [nine(16)] = '1' | y |
| nine_a_a_bradykinesia | radio | Were these changes meaningful to your quality of life? | 1, Yes \| 0, No \| 2, Don't know | [nine_9_bradykinesia] = '2' or [nine_9_bradykinesia] = '1' or [nine_9_bradykinesia] = '3' | y |
| nine_9_urinary | radio | To what extent do you feel that marijuana or cannabis improved the severity of urinary symptoms? | 1, Considerable improvement \| 2, Moderate improvement \| 3, Slight improvement \| 4, No improvement \| 5, Worsening of symptoms | [nine(17)] = '1' | y |
| nine_a_a_urinary | radio | Were these changes meaningful to your quality of life? | 1, Yes \| 0, No \| 2, Don't know | [nine_9_urinary] = '2' or [nine_9_urinary] = '1' or [nine_9_urinary] = '3' | y |
| nine_9_balance | radio | To what extent do you feel that marijuana or cannabis improved the severity of balance loss? | 1, Considerable improvement \| 2, Moderate improvement \| 3, Slight improvement \| 4, No improvement \| 5, Worsening of symptoms | [nine(18)] = '1' | y |
| nine_a_a_balance | radio | Were these changes meaningful to your quality of life? | 1, Yes \| 0, No \| 2, Don't know | [nine_9_balance] = '2' or [nine_9_balance] = '1' or [nine_9_balance] = '3' | y |
| nine_9_forgetfulness | radio | To what extent do you feel that marijuana or cannabis improved the severity of forgetfulness? | 1, Considerable improvement \| 2, Moderate improvement \| 3, Slight improvement \| 4, No improvement \| 5, Worsening of symptoms | [nine(19)] = '1' | y |
| nine_a_a_forgetfulness | radio | Were these changes meaningful to your quality of life? | 1, Yes \| 0, No \| 2, Don't know | [nine_9_forgetfulness] = '2' or [nine_9_forgetfulness] = '1' or [nine_9_forgetfulness] = '3' | y |
| nine_9_planning | radio | To what extent do you feel that marijuana or cannabis improved the severity of difficulty planning and/or multi-tasking? | 1, Considerable improvement \| 2, Moderate improvement \| 3, Slight improvement \| 4, No improvement \| 5, Worsening of symptoms | [nine(20)] = '1' | y |
| nine_a_a_planning | radio | Were these changes meaningful to your quality of life? | 1, Yes \| 0, No \| 2, Don't know | [nine_9_planning] = '2' or [nine_9_planning] = '1' or [nine_9_planning] = '3' | y |

| | | | | | |
|---|---|---|---|---|---|
| nine_9_hallucinations | radio | To what extent do you feel that marijuana or cannabis improved the severity of hallucinations? | 1, Considerable improvement \| 2, Moderate improvement \| 3, Slight improvement \| 4, No improvement \| 5, Worsening of symptoms | [nine(21)] = '1' | y |
| nine_a_a_hallucinations | radio | Were these changes meaningful to your quality of life? | 1, Yes \| 0, No \| 2, Don't know | [nine_9_hallucinations] = '2' or [nine_9_hallucinations] = '1' or [nine_9_hallucinations] = '3' | y |
| nine_9_other | radio | To what extent do you feel that marijuana or cannabis improved the severity of this other symptom? | 1, Considerable improvement \| 2, Moderate improvement \| 3, Slight improvement \| 4, No improvement \| 5, Worsening of symptoms | [nine(22)] = '1' | y |
| nine_a_a_other | radio | Were these changes meaningful to your quality of life? | 1, Yes \| 0, No \| 2, Don't know | [nine_9_other] = '2' or [nine_9_other] = '1' or [nine_9_other] = '3' | y |
| nine_b_appetite | radio | To what extent do you feel that marijuana or cannabis improved the frequency of appetite loss? | 1, Considerable improvement \| 2, Moderate improvement \| 3, Slight improvement \| 4, No improvement \| 5, Worsening of symptoms | [nine(1)] = '1' | y |
| nine_b_b_appetite | radio | Were these changes meaningful to your quality of life? | 1, Yes \| 0, No \| 2, Don't know | [nine_b_appetite] = '1' or [nine_b_appetite] = '2' or [nine_b_appetite] = '3' | y |
| nine_b_pai n | radio | To what extent do you feel that marijuana or cannabis improved the frequency of pain? | 1, Considerable improvement \| 2, Moderate improvement \| 3, Slight improvement \| 4, No improvement \| 5, Worsening of symptoms | [nine(2)] = '1' | y |
| nine_b_b_pain | radio | Were these changes meaningful to your quality of life? | 1, Yes \| 0, No \| 2, Don't know | [nine_b_pain] = '1' or [nine_b_pain] = '2' or [nine_b_pain] = '3' | y |
| nine_b_nausea | radio | To what extent do you feel that marijuana or cannabis improved the frequency of nausea? | 1, Considerable improvement \| 2, Moderate improvement \| 3, Slight improvement \| 4, No improvement \| 5, Worsening of symptoms | [nine(3)] = '1' | y |
| nine_b_b_nausea | radio | Were these changes meaningful to your quality of life? | 1, Yes \| 0, No \| 2, Don't know | [nine_b_nausea] = '1' or [nine_b_nausea] = '2' or [nine_b_nausea] = '3' | y |
| nine_b_anxiety | radio | To what extent do you feel that marijuana or cannabis improved the frequency of anxiety? | 1, Considerable improvement \| 2, Moderate improvement \| 3, Slight improvement \| 4, No improvement \| 5, Worsening of symptoms | [nine(4)] = '1' | y |
| nine_b_b_anxiety | radio | Were these changes meaningful to your quality of life? | 1, Yes \| 0, No \| 2, Don't know | [nine_b_anxiety] = '1' or [nine_b_anxiety] = '2' or [nine_b_anxiety] = '3' | y |
| nine_b_tremor | radio | To what extent do you feel that marijuana or cannabis | 1, Considerable improvement \| 2, Moderate improvement \| 3, Slight improvement \| 4, No improvement \| 5, Worsening of symptoms | [nine(5)] = '1' | y |

| | | | | | |
|---|---|---|---|---|---|
| | | improved the frequency of tremor? | | | |
| nine_b_b_tremor | radio | Were these changes meaningful to your quality of life? | 1, Yes \| 0, No \| 2, Don't know | [nine_b_tremor] = '1' or [nine_b_tremor] = '2' or [nine_b_tremor] = '3' | y |
| nine_b_stiffness | radio | To what extent do you feel that marijuana or cannabis improved the frequency of stiffness? | 1, Considerable improvement \| 2, Moderate improvement \| 3, Slight improvement \| 4, No improvement \| 5, Worsening of symptoms | [nine(6)] = '1' | y |
| nine_b_b_stiffness | radio | Were these changes meaningful to your quality of life? | 1, Yes \| 0, No \| 2, Don't know | [nine_b_stiffness] = '1' or [nine_b_stiffness] = '2' or [nine_b_stiffness] = '3' | y |
| nine_b_dyskinesias | radio | To what extent do you feel that marijuana or cannabis improved the frequency of dyskinesias? | 1, Considerable improvement \| 2, Moderate improvement \| 3, Slight improvement \| 4, No improvement \| 5, Worsening of symptoms | [nine(7)] = '1' | y |
| nine_b_b_dyskinesias | radio | Were these changes meaningful to your quality of life? | 1, Yes \| 0, No \| 2, Don't know | [nine_b_dyskinesias] = '1' or [nine_b_dyskinesias] = '2' or [nine_b_dyskinesias] = '3' | y |
| nine_b_depression | radio | To what extent do you feel that marijuana or cannabis improved the frequency of depression? | 1, Considerable improvement \| 2, Moderate improvement \| 3, Slight improvement \| 4, No improvement \| 5, Worsening of symptoms | [nine(8)] = '1' | y |
| nine_b_b_depression | radio | Were these changes meaningful to your quality of life? | 1, Yes \| 0, No \| 2, Don't know | [nine_b_depression] = '1' or [nine_b_depression] = '2' or [nine_b_depression] = '3' | y |
| nine_b_fatigue | radio | To what extent do you feel that marijuana or cannabis improved the frequency of fatigue? | 1, Considerable improvement \| 2, Moderate improvement \| 3, Slight improvement \| 4, No improvement \| 5, Worsening of symptoms | [nine(9)] = '1' | y |
| nine_b_b_fatigue | radio | Were these changes meaningful to your quality of life? | 1, Yes \| 0, No \| 2, Don't know | [nine_b_fatigue] = '1' or [nine_b_fatigue] = '2' or [nine_b_fatigue] = '3' | y |
| nine_b_freezing | radio | To what extent do you feel that marijuana or cannabis improved the frequency of freezing? | 1, Considerable improvement \| 2, Moderate improvement \| 3, Slight improvement \| 4, No improvement \| 5, Worsening of symptoms | [nine(10)] = '1' | y |
| nine_b_b_freezing | radio | Were these changes meaningful to your quality of life? | 1, Yes \| 0, No \| 2, Don't know | [nine_b_freezing] = '1' or [nine_b_freezing] = '2' or [nine_b_freezing] = '3' | y |
| nine_b_constipation | radio | To what extent do you feel that marijuana or cannabis improved the frequency of constipation? | 1, Considerable improvement \| 2, Moderate improvement \| 3, Slight improvement \| 4, No improvement \| 5, Worsening of symptoms | [nine(11)] = '1' | y |

| | | | | | |
|---|---|---|---|---|---|
| nine_b_b_constipation | radio | Were these changes meaningful to your quality of life? | 1, Yes \| 0, No \| 2, Don't know | [nine_b_constipation] = '1' or [nine_b_constipation] = '2' or [nine_b_constipation] = '3' | y |
| nine_b_motivation | radio | To what extent do you feel that marijuana or cannabis improved the frequency of motivation loss? | 1, Considerable improvement \| 2, Moderate improvement \| 3, Slight improvement \| 4, No improvement \| 5, Worsening of symptoms | [nine(12)] = '1' | y |
| nine_b_b_motivation | radio | Were these changes meaningful to your quality of life? | 1, Yes \| 0, No \| 2, Don't know | [nine_b_motivation] = '1' or [nine_b_motivation] = '2' or [nine_b_motivation] = '3' | y |
| nine_b_sleepiness | radio | To what extent do you feel that marijuana or cannabis improved the frequency of sleepiness (daytime)? | 1, Considerable improvement \| 2, Moderate improvement \| 3, Slight improvement \| 4, No improvement \| 5, Worsening of symptoms | [nine(13)] = '1' | y |
| nine_b_b_sleepiness | radio | Were these changes meaningful to your quality of life? | 1, Yes \| 0, No \| 2, Don't know | [nine_b_sleepiness] = '1' or [nine_b_sleepiness] = '2' or [nine_b_sleepiness] = '3' | y |
| nine_b_sleepdisorders | radio | To what extent do you feel that marijuana or cannabis improved the frequency of sleep disorders? | 1, Considerable improvement \| 2, Moderate improvement \| 3, Slight improvement \| 4, No improvement \| 5, Worsening of symptoms | [nine(14)] = '1' | y |
| nine_b_b_sleepdisorders | radio | Were these changes meaningful to your quality of life? | 1, Yes \| 0, No \| 2, Don't know | [nine_b_sleepdisorders] = '1' or [nine_b_sleepdisorders] = '2' or [nine_b_sleepdisorders] = '3' | y |
| nine_b_dystonia | radio | To what extent do you feel that marijuana or cannabis improved the frequency of dystonia? | 1, Considerable improvement \| 2, Moderate improvement \| 3, Slight improvement \| 4, No improvement \| 5, Worsening of symptoms | [nine(15)] = '1' | y |
| nine_b_b_dystonia | radio | Were these changes meaningful to your quality of life? | 1, Yes \| 0, No \| 2, Don't know | [nine_b_dystonia] = '1' or [nine_b_dystonia] = '2' or [nine_b_dystonia] = '3' | y |
| nine_b_bradykinesia | radio | To what extent do you feel that marijuana or cannabis improved the frequency of bradykinesia (slowness of movement)? | 1, Considerable improvement \| 2, Moderate improvement \| 3, Slight improvement \| 4, No improvement \| 5, Worsening of symptoms | [nine(16)] = '1' | y |
| nine_b_b_bradykinesia | radio | Were these changes meaningful to your quality of life? | 1, Yes \| 0, No \| 2, Don't know | [nine_b_bradykinesia] = '1' or [nine_b_bradykinesia] = '2' or [nine_b_bradykinesia] = '3' | y |

| | | | | | |
|---|---|---|---|---|---|
| nine_b_urinary | radio | To what extent do you feel that marijuana or cannabis improved the frequency of urinary symptoms? | 1, Considerable improvement \| 2, Moderate improvement \| 3, Slight improvement \| 4, No improvement \| 5, Worsening of symptoms | [nine(17)] = '1' | y |
| nine_b_b_urinary | radio | Were these changes meaningful to your quality of life? | 1, Yes \| 0, No \| 2, Don't know | [nine_b_urinary] = '1' or [nine_b_urinary] = '2' or [nine_b_urinary] = '3' | y |
| nine_b_balance | radio | To what extent do you feel that marijuana or cannabis improved the frequency of balance loss? | 1, Considerable improvement \| 2, Moderate improvement \| 3, Slight improvement \| 4, No improvement \| 5, Worsening of symptoms | [nine(18)] = '1' | y |
| nine_b_b_balance | radio | Were these changes meaningful to your quality of life? | 1, Yes \| 0, No \| 2, Don't know | [nine_b_balance] = '1' or [nine_b_balance] = '2' or [nine_b_balance] = '3' | y |
| nine_b_forgetfulness | radio | To what extent do you feel that marijuana or cannabis improved the frequency of forgetfulness? | 1, Considerable improvement \| 2, Moderate improvement \| 3, Slight improvement \| 4, No improvement \| 5, Worsening of symptoms | [nine(19)] = '1' | y |
| nine_b_b_forgetfulness | radio | Were these changes meaningful to your quality of life? | 1, Yes \| 0, No \| 2, Don't know | [nine_b_forgetfulness] = '1' or [nine_b_forgetfulness] = '2' or [nine_b_forgetfulness] = '3' | y |
| nine_b_planning | radio | To what extent do you feel that marijuana or cannabis improved the frequency of difficulty planning and/or multi-tasking? | 1, Considerable improvement \| 2, Moderate improvement \| 3, Slight improvement \| 4, No improvement \| 5, Worsening of symptoms | [nine(20)] = '1' | y |
| nine_b_b_planning | radio | Were these changes meaningful to your quality of life? | 1, Yes \| 0, No \| 2, Don't know | [nine_b_planning] = '1' or [nine_b_planning] = '2' or [nine_b_planning] = '3' | y |
| nine_b_hallucinations | radio | To what extent do you feel that marijuana or cannabis improved the frequency of hallucinations? | 1, Considerable improvement \| 2, Moderate improvement \| 3, Slight improvement \| 4, No improvement \| 5, Worsening of symptoms | [nine(21)] = '1' | y |
| nine_b_b_hallucinations | radio | Were these changes meaningful to your quality of life? | 1, Yes \| 0, No \| 2, Don't know | [nine_b_hallucinations] = '1' or [nine_b_hallucinations] = '2' or [nine_b_hallucinations] = '3' | y |
| nine_b_other | radio | To what extent do you feel that marijuana or cannabis improved the frequency of this other symptom? | 1, Considerable improvement \| 2, Moderate improvement \| 3, Slight improvement \| 4, No improvement \| 5, Worsening of symptoms | [nine(22)] = '1' | y |

| | | | | | |
|---|---|---|---|---|---|
| nine_b_b_other | radio | Were these changes meaningful to your quality of life? | 1, Yes \| 0, No \| 2, Don't know | [nine_b_other] = '1' or [nine_b_other] = '2' or [nine_b_other] = '3' | y |
| ten | checkbox | For which of the following medication side effects do you use marijuana or cannabis to decrease? Select all that apply. | 1, Anxiety \| 2, Nausea/ Vomiting \| 3, Lightheadedness \| 4, Loss of appetite \| 5, Low blood pressure \| 6, Confusion \| 7, Hallucinations \| 8, Dyskinesia \| 9, Gastrointestinal issues \| 10, My medication side effect is not listed on this list | [two_d] = '3' | y |
| ten_other | text | For what other medication side effect do you use marijuana or cannabis to decrease? | | [ten(10)] = '1' | y |
| ten_a_anxiety | radio | To what extent do you feel that marijuana or cannabis decreased the severity of anxiety? | 1, Considerable decrease \| 2, Moderate decrease \| 3, Slight decrease \| 4, No decrease \| 5, Worsening of side effects | [ten(1)] = '1' | y |
| ten_a_a_anxiety | radio | Were these changes meaningful to your quality of life? | 1, Yes \| 0, No \| 2, Don't know | [ten_a_anxiety] = '1' or [ten_a_anxiety] = '2' or [ten_a_anxiety] = '3' | y |
| ten_a_nausea | radio | To what extent do you feel that marijuana or cannabis decreased the severity of nausea/vomiting? | 1, Considerable decrease \| 2, Moderate decrease \| 3, Slight decrease \| 4, No decrease \| 5, Worsening of side effects | [ten(2)] = '1' | y |
| ten_a_a_nausea | radio | Were these changes meaningful to your quality of life? | 1, Yes \| 0, No \| 2, Don't know | [ten_a_nausea] = '1' or [ten_a_nausea] = '2' or [ten_a_nausea] = '3' | y |
| ten_a_lightheadedness | radio | To what extent do you feel that marijuana or cannabis decreased the severity of lightheadedness? | 1, Considerable decrease \| 2, Moderate decrease \| 3, Slight decrease \| 4, No decrease \| 5, Worsening of side effects | [ten(3)] = '1' | y |
| ten_a_a_lightheadedness | radio | Were these changes meaningful to your quality of life? | 1, Yes \| 0, No \| 2, Don't know | [ten_a_lightheadedness] = '1' or [ten_a_lightheadedness] = '2' or [ten_a_lightheadedness] = '3' | y |
| ten_a_appetite | radio | To what extent do you feel that marijuana or cannabis decreased the severity of loss of appetite? | 1, Considerable decrease \| 2, Moderate decrease \| 3, Slight decrease \| 4, No decrease \| 5, Worsening of side effects | [ten(4)] = '1' | y |
| ten_a_a_appetite | radio | Were these changes meaningful to your quality of life? | 1, Yes \| 0, No \| 2, Don't know | [ten_a_appetite] = '1' or [ten_a_appetite] = '2' or [ten_a_appetite] = '3' | y |
| ten_a_bloodpressure | radio | To what extent do you feel that marijuana or cannabis decreased the severity of low blood pressure? | 1, Considerable decrease \| 2, Moderate decrease \| 3, Slight decrease \| 4, No decrease \| 5, Worsening of side effects | [ten(5)] = '1' | y |
| ten_a_a_bloodpressure | radio | Were these changes meaningful to your quality of life? | 1, Yes \| 0, No \| 2, Don't know | [ten_a_bloodpressure] = '1' or [ten_a_bloodpressure] = | y |

| | | | | | |
|---|---|---|---|---|---|
| | | | | '2' or [ten_a_bloodpressure] = '3' | |
| ten_a_confusion | radio | To what extent do you feel that marijuana or cannabis decreased the severity of confusion? | 1, Considerable decrease \| 2, Moderate decrease \| 3, Slight decrease \| 4, No decrease \| 5, Worsening of side effects | [ten(6)] = '1' | y |
| ten_a_a_confusion | radio | Were these changes meaningful to your quality of life? | 1, Yes \| 0, No \| 2, Don't know | [ten_a_confusion] = '1' or [ten_a_confusion] = '2' or [ten_a_confusion] = '3' | y |
| ten_a_hallucinations | radio | To what extent do you feel that marijuana or cannabis decreased the severity of hallucinations? | 1, Considerable decrease \| 2, Moderate decrease \| 3, Slight decrease \| 4, No decrease \| 5, Worsening of side effects | [ten(7)] = '1' | y |
| ten_a_a_hallucinations | radio | Were these changes meaningful to your quality of life? | 1, Yes \| 0, No \| 2, Don't know | [ten_a_hallucinations] = '1' or [ten_a_hallucinations] = '2' or [ten_a_hallucinations] = '3' | y |
| ten_a_dyskinesia | radio | To what extent do you feel that marijuana or cannabis decreased the severity of dyskinesia? | 1, Considerable decrease \| 2, Moderate decrease \| 3, Slight decrease \| 4, No decrease \| 5, Worsening of side effects | [ten(8)] = '1' | y |
| ten_a_a_dyskinesia | radio | Were these changes meaningful to your quality of life? | 1, Yes \| 0, No \| 2, Don't know | [ten_a_dyskinesia] = '1' or [ten_a_dyskinesia] = '2' or [ten_a_dyskinesia] = '3' | y |
| ten_a_gi | radio | To what extent do you feel that marijuana or cannabis decreased the severity of gastrointestinal issues? | 1, Considerable decrease \| 2, Moderate decrease \| 3, Slight decrease \| 4, No decrease \| 5, Worsening of side effects | [ten(9)] = '1' | y |
| ten_a_a_gi | radio | Were these changes meaningful to your quality of life? | 1, Yes \| 0, No \| 2, Don't know | [ten_a_gi] = '1' or [ten_a_gi] = '2' or [ten_a_gi] = '3' | y |
| ten_a_other | radio | To what extent do you feel that marijuana or cannabis decreased the severity of this other side effect? | 1, Considerable decrease \| 2, Moderate decrease \| 3, Slight decrease \| 4, No decrease \| 5, Worsening of side effects | [ten(10)] = '1' | y |
| ten_a_a_gi_2 | radio | Were these changes meaningful to your quality of life? | 1, Yes \| 0, No \| 2, Don't know | [ten_a_other] = '1' or [ten_a_other] = '2' or [ten_a_other] = '3' | y |
| ten_b_anxiety | radio | To what extent do you feel that marijuana or cannabis decreased the frequency of anxiety? | 1, Considerable decrease \| 2, Moderate decrease \| 3, Slight decrease \| 4, No decrease \| 5, Worsening of side effects | [ten(1)] = '1' | y |
| ten_b_b_anxiety | radio | Were these changes meaningful to your quality of life? | 1, Yes \| 0, No \| 2, Don't know | [ten_b_anxiety] = '1' or [ten_b_anxiety] = '2' or [ten_b_anxiety] = '3' | y |

| | | | | | |
|---|---|---|---|---|---|
| ten_b_other | radio | To what extent do you feel that marijuana or cannabis decreased the frequency of this other side effect? | 1, Considerable decrease \| 2, Moderate decrease \| 3, Slight decrease \| 4, No decrease \| 5, Worsening of side effects | [ten(10)] = '1' | y |
| ten_b_b_anxiety_2 | radio | Were these changes meaningful to your quality of life? | 1, Yes \| 0, No \| 2, Don't know | [ten_b_other] = '1' or [ten_b_other] = '2' or [ten_b_other] = '3' | y |
| ten_b_nausea | radio | To what extent do you feel that marijuana or cannabis decreased the frequency of nausea/vomiting? | 1, Considerable decrease \| 2, Moderate decrease \| 3, Slight decrease \| 4, No decrease \| 5, Worsening of side effects | [ten(2)] = '1' | y |
| ten_b_b_nausea | radio | Were these changes meaningful to your quality of life? | 1, Yes \| 0, No \| 2, Don't know | [ten_b_nausea] = '1' or [ten_b_nausea] = '2' or [ten_b_nausea] = '3' | y |
| ten_b_lightheaded | radio | To what extent do you feel that marijuana or cannabis decreased the frequency of lightheadedness? | 1, Considerable decrease \| 2, Moderate decrease \| 3, Slight decrease \| 4, No decrease \| 5, Worsening of side effects | [ten(3)] = '1' | y |
| ten_b_b_lightheaded | radio | Were these changes meaningful to your quality of life? | 1, Yes \| 0, No \| 2, Don't know | [ten_b_lightheaded] = '1' or [ten_b_lightheaded] = '2' or [ten_b_lightheaded] = '3' | y |
| ten_b_appetite | radio | To what extent do you feel that marijuana or cannabis decreased the frequency of loss of appetite? | 1, Considerable decrease \| 2, Moderate decrease \| 3, Slight decrease \| 4, No decrease \| 5, Worsening of side effects | [ten(4)] = '1' | y |
| ten_b_b_appetite | radio | Were these changes meaningful to your quality of life? | 1, Yes \| 0, No \| 2, Don't know | [ten_b_appetite] = '1' or [ten_b_appetite] = '2' or [ten_b_appetite] = '3' | y |
| ten_b_bloodpressure | radio | To what extent do you feel that marijuana or cannabis decreased the frequency of low blood pressure? | 1, Considerable decrease \| 2, Moderate decrease \| 3, Slight decrease \| 4, No decrease \| 5, Worsening of side effects | [ten(5)] = '1' | y |
| ten_b_b_bloodpressure | radio | Were these changes meaningful to your quality of life? | 1, Yes \| 0, No \| 2, Don't know | [ten_b_bloodpressure] = '1' or [ten_b_bloodpressure] = '2' or [ten_b_bloodpressure] = '3' | y |
| ten_b_confusion | radio | To what extent do you feel that marijuana or cannabis decreased the frequency of confusion? | 1, Considerable decrease \| 2, Moderate decrease \| 3, Slight decrease \| 4, No decrease \| 5, Worsening of side effects | [ten(6)] = '1' | y |
| ten_b_b_confusion | radio | Were these changes meaningful to your quality of life? | 1, Yes \| 0, No \| 2, Don't know | [ten_b_confusion] = '1' or [ten_b_confusion] = '2' or [ten_b_confusion] = '3' | y |

| | | | | | | |
|---|---|---|---|---|---|---|
| ten_b_hallucinations | | radio | To what extent do you feel that marijuana or cannabis decreased the frequency of hallucinations? | 1, Considerable decrease \| 2, Moderate decrease \| 3, Slight decrease \| 4, No decrease \| 5, Worsening of side effects | [ten(7)] = '1' | y |
| ten_b_b_hallucinations | | radio | Were these changes meaningful to your quality of life? | 1, Yes \| 0, No \| 2, Don't know | [ten_b_hallucinations] = '1' or [ten_b_hallucinations] = '2' or [ten_b_hallucinations] = '3' | y |
| ten_b_diskynesia | | radio | To what extent do you feel that marijuana or cannabis decreased the frequency of dyskinesia? | 1, Considerable decrease \| 2, Moderate decrease \| 3, Slight decrease \| 4, No decrease \| 5, Worsening of side effects | [ten(8)] = '1' | y |
| ten_b_b_diskynesia | | radio | Were these changes meaningful to your quality of life? | 1, Yes \| 0, No \| 2, Don't know | [ten_b_diskynesia] = '1' or [ten_b_diskynesia] = '2' or [ten_b_diskynesia] = '3' | y |
| ten_b_gi | | radio | To what extent do you feel that marijuana or cannabis decreased the frequency of gastrointestinal issues? | 1, Considerable decrease \| 2, Moderate decrease \| 3, Slight decrease \| 4, No decrease \| 5, Worsening of side effects | [ten(9)] = '1' | y |
| ten_b_b_gi | | radio | Were these changes meaningful to your quality of life? | 1, Yes \| 0, No \| 2, Don't know | [ten_b_gi] = '1' or [ten_b_gi] = '2' or [ten_b_gi] = '3' | y |
| eleven | | radio | Do you feel that marijuana or cannabis adequately address your PD motor symptoms? | 1, Completely address \| 2, Mostly address \| 3, Somewhat address \| 4, Do not address | [pwp] = '1' and [one] = '1' | y |
| twelve | | radio | Do you feel that marijuana or cannabis adequately address your PD non-motor symptoms? | 1, Completely address \| 2, Mostly address \| 3, Somewhat address \| 4, Do not address | [pwp] = '1' and [one] = '1' | y |
| thirteen | | radio | Have you experienced any negative side effects to marijuana or cannabis use? | 1, Yes \| 0, No | ([pwp] = '1' and [one] = '1') or ([pwp] = '1' and [one_a(5)] = '1') | y |
| thirteen_a | | checkbox | What side effects did you experience when using marijuana or cannabis? Select all that apply. | 1, Anxiety/paranoia \| 2, Hallucinations/psychosis \| 3, Loss of motivation/ apathy \| 4, Depression \| 5, Forgetfulness \| 6, Cough \| 7, Nausea \| 8, Stomach pain \| 9, Weight gain \| 10, Increased heart rate \| 11, Impaired coordination \| 12, Loss of balance/falls \| 13, Dizziness \| 14, A side effect that is not listed on this list | [thirteen] = '1' | y |
| thirteen_a_a | | text | What side effect did you experience that was not on the list? | | [thirteen_a(14)] = '1' | y |
| fourteen | | notes | Other than side effects, have you experienced any negative consequences to marijuana or cannabis use? | | [thirteen] = '1' | y |
| fifteen | | radio | In the past six months, have you stopped using marijuana or cannabis? | 1, Yes \| 0, No | [pwp] = '1' and [one] = '1' | y |

| | | | | | | |
|---|---|---|---|---|---|---|
| fifteen_a | | checkbox | For what reason did you stop using marijuana or cannabis? Select all that apply. | 1, My symptoms did not improve while using marijuana or cannabis \| 2, My symptoms became worse while using marijuana or cannabis \| 3, My prescription medication side effects did not improve while using marijuana or cannabis \| 4, I changed my prescription medications \| 5, I experienced unwanted side effects while using marijuana or cannabis \| 6, The cost of marijuana or cannabis was too expensive \| 7, I was unable to access or acquire marijuana or cannabis \| 8, I was uncomfortable with the stigma of using marijuana or cannabis \| 9, I felt that I was developing a dependency to marijuana or cannabis \| 10, My health team discouraged me from using marijuana or cannabis \| 11, Other | [fifteen] = '1' or ([pwp] = '1' and [one_a(5)] = '1') | y |
| fifteen_a_a | | notes | For what reason other did you stop using marijuana or cannabis? | | [fifteen_a(11)] = '1' | y |
| sixteen | | notes | What does using marijuana or cannabis allow you to do that you could not do otherwise? | | [fifteen] = '0' | y |
| seventeen | | notes | What were your reasons for trying marijuana or cannabis to treat your Parkinson's disease over other alternative therapies? | | [pwp] = '1' and [one] = '1' and ([two] = '1' or [two] = '2' or [two] = '4') | y |

| | | | | | | |
|---|---|---|---|---|---|---|
| eighteen | Prescription Medication Satisfaction | radio | Do you feel that your prescribed PD medications (i.e. Sinemet, Mirapex) adequately address the motor aspects of your Parkinson's disease? | 1, Completely address \| 2, Mostly address \| 3, Somewhat address \| 4, Do not address \| 0, Not currently taking medications for PD | [pwp] = '1' | y |
| nineteen | | radio | Do you feel that your prescription medications (PD medications or other prescription medications) adequately address the non-motor aspects of your Parkinson's disease? | 1, Completely address \| 2, Mostly address \| 3, Somewhat address \| 4, Do not address \| 0, Not currently taking medications for PD | [pwp] = '1' | y |
| nineteen_a | | radio | Were you concerned about potential interactions between your prescribed PD medications and marijuana or cannabis? | 1, Not concerned \| 2, Somewhat concerned \| 3, Very concerned | [one] = '1' and ([eighteen] = '1' or [eighteen] = '2' or [eighteen] = '3' or [eighteen] = '4') | y |
| twenty | | radio | When you began using marijuana or cannabis, did you think that it would be a replacement for PD prescription drugs? | 1, Yes \| 0, No | [pwp] = '1' and [one] = '1' and ([two] = '1' or [two] = '2' or [two] = '4') | y |
| twentyone | | radio | Has your experience with marijuana or cannabis | 1, Eliminated the use for PD prescription drugs \| 2, Significantly reduced the use of PD prescription drugs \| 3, Reduced the use of PD prescription drugs \| | [pwp] = '1' and [one] = '1' | y |

| | | | | | | |
|---|---|---|---|---|---|---|
| | | | impacted your use of PD prescription drugs? | 4, Has had no impact on the use of PD prescription drugs \| 5, Increased the use of PD prescription drugs | | |
| twentytwo | Clinical Trial Interest | radio | Would you be interested in learning more information about a clinical trial exploring the impact of marijuana or cannabis on Parkinson's disease symptoms? | 1, Yes \| 0, No | [pwp] = '1' | y |
| twentythree | | radio | Would you be interested in enrolling in a clinical trial exploring the impact of marijuana or cannabis on Parkinson's disease symptoms? | 1, Yes \| 0, No | [pwp] = '1' | y |
| twentyfour | | radio | Have you previously participated in a survey about cannabis usage for Fox Insight? | 1, Yes \| 0, No | [pwp] = '1' | y |
| b | Demographic Information | dropdown | In what year were you born? | 1919, Prior to 1920 \| 1920, 1920 \| 1921, 1921 \| 1922, 1922 \| 1923, 1923 \| 1924, 1924 \| 1925, 1925 \| 1926, 1926 \| 1927, 1927 \| 1928, 1928 \| 1929, 1929 \| 1930, 1930 \| 1931, 1931 \| 1932, 1932 \| 1933, 1933 \| 1934, 1934 \| 1935, 1935 \| 1936, 1936 \| 1937, 1937 \| 1938, 1938 \| 1939, 1939 \| 1940, 1940 \| 1941, 1941 \| 1942, 1942 \| 1943, 1943 \| 1944, 1944 \| 1945, 1945 \| 1946, 1946 \| 1947, 1947 \| 1948, 1948 \| 1949, 1949 \| 1950, 1950 \| 1951, 1951 \| 1952, 1952 \| 1953, 1953 \| 1954, 1954 \| 1955, 1955 \| 1956, 1956 \| 1957, 1957 \| 1958, 1958 \| 1959, 1959 \| 1960, 1960 \| 1961, 1961 \| 1962, 1962 \| 1963, 1963 \| 1964, 1964 \| 1965, 1965 \| 1966, 1966 \| 1967, 1967 \| 1968, 1968 \| 1969, 1969 \| 1970, 1970 \| 1971, 1971 \| 1972, 1972 \| 1973, 1973 \| 1974, 1974 \| 1975, 1975 \| 1976, 1976 \| 1977, 1977 \| 1978, 1978 \| 1979, 1979 \| 1980, 1980 \| 1981, 1981 \| 1982, 1982 \| 1983, 1983 \| 1984, 1984 \| 1985, 1985 \| 1986, 1986 \| 1987, 1987 \| 1988, 1988 \| 1989, 1989 \| 1990, 1990 \| 1991, 1991 \| 1992, 1992 \| 1993, 1993 \| 1994, 1994 \| 1995, 1995 \| 1996, 1996 \| 1997, 1997 \| 1998, 1998 \| 1999, 1999 \| 2000, 2000 \| 2001, 2001 \| 2002, 2002 \| 2003, 2003 \| 2004, 2004 \| 2005, 2005 \| 2006, 2006 \| 2007, 2007 \| 2008, 2008 \| 2009, 2009 \| 2010, 2010 \| 2011, 2011 \| 2012, 2012 \| 2013, 2013 \| 2014, 2014 \| 2015, 2015 \| 2016, 2016 \| 2017, 2017 \| 2018, 2018 \| 2019, 2019 | [pwp] = '1' | y |
| c | | dropdown | In what year were you diagnosed Parkinson's disease? | 1939, Prior to 1940 \| 1940, 1940 \| 1941, 1941 \| 1942, 1942 \| 1943, 1943 \| 1944, 1944 \| 1945, 1945 \| 1946, 1946 \| 1947, 1947 \| 1948, 1948 \| 1949, 1949 \| 1950, | [pwp] = '1' | y |

| | | | | | | |
|---|---|---|---|---|---|---|
| | | | | 1950 \| 1951, 1951 \| 1952, 1952 \| 1953, 1953 \| 1954, 1954 \| 1955, 1955 \| 1956, 1956 \| 1957, 1957 \| 1958, 1958 \| 1959, 1959 \| 1960, 1960 \| 1961, 1961 \| 1962, 1962 \| 1963, 1963 \| 1964, 1964 \| 1965, 1965 \| 1966, 1966 \| 1967, 1967 \| 1968, 1968 \| 1969, 1969 \| 1970, 1970 \| 1971, 1971 \| 1972, 1972 \| 1973, 1973 \| 1974, 1974 \| 1975, 1975 \| 1976, 1976 \| 1977, 1977 \| 1978, 1978 \| 1979, 1979 \| 1980, 1980 \| 1981, 1981 \| 1982, 1982 \| 1983, 1983 \| 1984, 1984 \| 1985, 1985 \| 1986, 1986 \| 1987, 1987 \| 1988, 1988 \| 1989, 1989 \| 1990, 1990 \| 1991, 1991 \| 1992, 1992 \| 1993, 1993 \| 1994, 1994 \| 1995, 1995 \| 1996, 1996 \| 1997, 1997 \| 1998, 1998 \| 1999, 1999 \| 2000, 2000 \| 2001, 2001 \| 2002, 2002 \| 2003, 2003 \| 2004, 2004 \| 2005, 2005 \| 2006, 2006 \| 2007, 2007 \| 2008, 2008 \| 2009, 2009 \| 2010, 2010 \| 2011, 2011 \| 2012, 2012 \| 2013, 2013 \| 2014, 2014 \| 2015, 2015 \| 2016, 2016 \| 2017, 2017 \| 2018, 2018 \| 2019, 2019 \| 2020, 2020 | | |
| d | | radio | Is Parkinson's disease your primary health concern? | 1, Yes \| 0, No | [pwp] = '1' | y |
| e | | radio | Do you have a care partner? | 1, Yes \| 0, No | [pwp] = '1' | y |
| f | | radio | What is your marital status? | 1, Single (never married) \| 2, Married or domestic partnership \| 3, Separated \| 4, Widowed \| 5, Divorced | [pwp] = '1' | y |
| g | | radio | Please select the gender you identify with: | 1, Male \| 2, Female \| 3, Other \| 4, Prefer to not answer | [pwp] = '1' | y |
| h | | radio | Please select the category that describes you. | 1, American Indian or Alaska Native \| 2, Asian \| 3, Black/African American \| 4, Hispanic/Latino \| 5, Native Hawaiian Pacific Islander \| 6, White \| 7, Mixed \| 8, Some other race, ethnicity or origin \| 9, Prefer to not answer | [pwp] = '1' | y |
| h_a | | text | What other race, ethnicity or origin do you identify with? | | [h] = '8' | y |
| i | | radio | What is your current employment status? | 1, Employed full time \| 2, Employed part time \| 3, Self-employed \| 4, Unemployed \| 5, Retired \| 6, Unable to work | [pwp] = '1' | y |
| j | | radio | What is your highest level of education? | 1, Less than a high school diploma \| 2, High school degree or equivalent (e.g. GED) \| 3, Some college, no degree \| 4, Trade/technical school (vocational training) \| 5, Associate degree (e.g. AA, AS) \| 6, Bachelor's degree (e.g. BA, BS) \| 7, Master's degree (e.g. MA, MS, MEd) \| 8, Professional degree (e.g. MD, DDS, DVM) \| 9, Doctorate (e.g. PhD, EdD) | [pwp] = '1' | y |
| k | | radio | What is your current yearly household income? (include income from | 1, Less than $25,000 \| 2, $25,000 to $34,999 \| 3, $35,000 to $49,999 \| 4, $50,000 to $74,999 \| 5, $75,000 to $99,999 \| 6, $100,000 to $149,999 \| 7, | [pwp] = '1' | y |

| | | | | | | |
|---|---|---|---|---|---|---|
| | | | Social Security, investment, etc.): | $150,000 to $199,999 \| 8, $200,000 or more \| 9, Prefer not to say | | |
| I | | text | Please enter the first three numbers of your zip code. For example, if your zip code is 07042, please enter 070. | | [pwp] = '1' | y |
